# Integrating Quantitative Histology with Clinical Data Improves Prediction of Cervical Intraepithelial Neoplasia Regression

**DOI:** 10.64898/2026.01.21.26344510

**Authors:** Oskari Lehtonen, Niko Nordlund, Essi Kahelin, Laura Bergqvist, Karoliina Aro, Sampsa Hautaniemi, Ilkka Kalliala, Anni Virtanen

## Abstract

Cervical intraepithelial neoplasia grade 2 (CIN2) lesions show variable outcomes, and accurate prediction of regression remains a major clinical challenge. We developed an interpretable machine learning pipeline that integrates quantitative histological, clinical, and human papillomavirus (HPV) -genotyping data to predict lesion regression within one and two years. Using panoptic segmentation of routine hematoxylin and eosin (H&E) -stained biopsies, we extracted human-interpretable morphological and immune cell infiltration related features that capture the key histopathological characteristics of CIN2 and identified features that predicted lesion regression. Further, integrating these features to predictive clinical features achieved higher predictive accuracy than clinical variables alone. These findings demonstrate that quantitative, interpretable analysis of H&E histology of routine diagnostic biopsies contains relevant information that predicts the natural history of CIN2 lesions.

**Graphical Abstract:** Created in BioRender. Lehtonen, O. (2026) https://BioRender.com/rlnkbkp

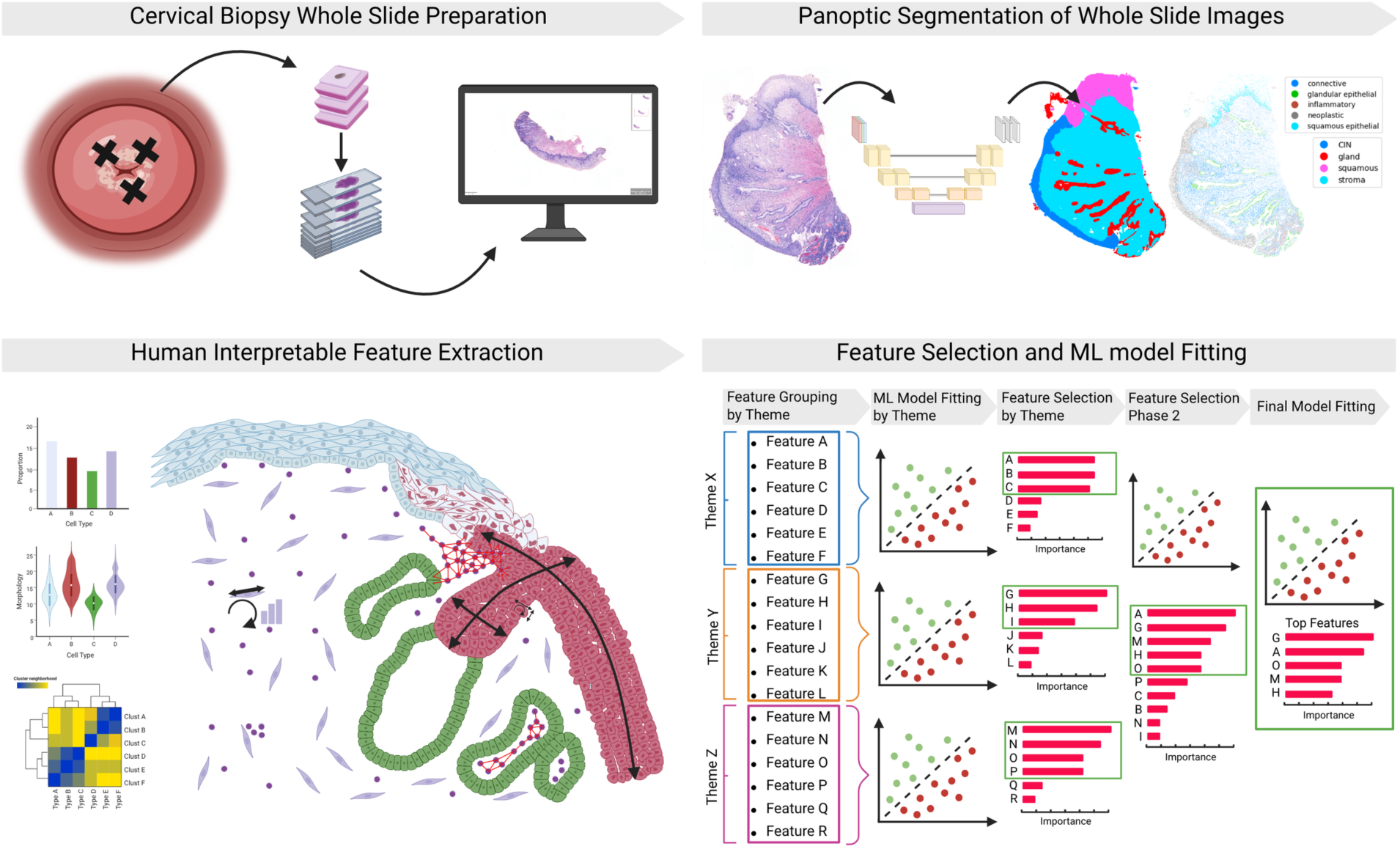

## Introduction

Cervical cancer develops from persistent human papillomavirus (HPV) infection through detectable precursor lesions that can be treated before progression into invasive cancer [1]. The histopathological cancer precursor of squamous cervical cancer, high-grade squamous intraepithelial lesion (HSIL), can be further divided into two lesion types, cervical intraepithelial neoplasia (CIN) grade 2 and 3, according to the extent of the dysplastic cells in the tissue, i.e. severity of the lesion. CIN3 carries a significant risk of invasive cancer and is always treated [2]. CIN2 lesions, however, can progress, persist or regress spontaneously. In a systematic review and meta-analysis approximately 50% of CIN2 lesions regressed spontaneously within 24 months of follow-up [3]. The option of active surveillance of HSIL/CIN2 instead of immediate treatment is included in many treatment guidelines [4].

Still, the natural progression of CIN2 lesions is unpredictable. It is influenced by the interplay of viral (HPV) factors, host factors, and other factors, such as the microbiome [5]. Very few biomarkers on whether a CIN2-lesion would spontaneously regress and could hence be managed with follow-up exist. Further, even though every CIN2 diagnosis is based on the morphology of the lesion in histological slides, there are no histology-based biomarkers or features that would predict regression potential.

We hypothesize that the histology of the lesion in the diagnostic biopsies contain information enabling the estimation of regression potential. Viral replication-related and transformative changes in cell and tissue morphology and respective microenvironmental responses are visible in the routine hematoxylin and eosin (H&E) stained histology of tissue biopsies [6]. While visual assessment of such features is subjective and labor-intensive, recent studies in other cancer types have shown that quantitatively extracted, interpretable histological features derived using deep learning (DL)–based segmentation and image analysis techniques can be used by machine learning (ML) methods to systematically identify the most informative predictors [7–9].

To discover predictive histological markers for CIN2 lesion regression, we analyzed routine diagnostic H&E-stained biopsies from a cohort of women participating in a prospective active surveillance study [10]. We used DL and image analysis to objectively extract histological features, which were then combined with clinical and HPV genotyping data to identify predictors of regression potential. In addition to aiming at predicting the regression of the lesions within the two year follow-up of the study protocol, we were especially interested in predicting which lesions will regress quickly (within a year), as among these women active surveillance instead of immediate treatment would be most beneficial both to the patient and in terms of used health care resources.

## Materials and Methods

### Cohort Description

Our cohort consisted of 146 women under 31 years who were diagnosed with CIN2 and enrolled in a study of active surveillance rather than immediate treatment at the Helsinki University Hospital [10]. In this study, regression was defined as achieving normal histological and cytological results at two consecutive visits within the 2–3-year follow-up period (for women with normal results at 2 years, also the following samples at 3 years were considered) and quick regression as normal histological and cytological results at two consecutive visits within the first year from diagnosis. The rest of the cases, i.e. cases that showed signs of progression requiring loop electrosurgical excision procedure (LEEP) within 2 years, lesion persistence, or only partial regression (low-grade cytological/histological findings) at the end of 2-year follow-up were classified as non-regressing. With these criteria, 38 (26.0 %) out of the 146 patients regressed within one year and were defined as quick regressing. Altogether 88 (60.3%) patients regressed within 2-3 years, and 58 (39.7%) were defined as non-regressing.

H&E-stained slides from all diagnostic biopsies with CIN2 were scanned at 20x magnification with Pannoramic P250 scanner at the Helsinki biobank; the scans were visually evaluated for tissue quality and slides with extreme tissue fragmentation were excluded. Eventually, 185 whole slide images (WSIs) were included in the analysis. Immunohistochemical staining for p16-INK4A was performed on 173 out of the 185 biopsies to confirm that the lesions were high-risk HPV related.

### Panoptic Segmentation of H&E-Stained WSIs

To enable extraction of histological features from H&E-stained CIN2 biopsy WSIs, panoptic segmentation was applied to delineate nuclear and tissue region objects in the WSIs [11]. We used the Histolytics library (v0.2.2) implementation of the Cellpose model, pre-trained on histological cervical data (https://huggingface.co/histolytics-hub/cellpose-histo-cin2-pan-v1) [12,13]. The WSI-level panoptic segmentation maps were visually inspected using QuPath (v0.5.0) [14], and any clear segmentation errors, such as misclassification of healthy basal squamous epithelium as neoplastic lesion, were manually corrected to ensure accurate downstream feature extraction.

### Histological Features

Using the *Histolytics* (v0.2.2) library, we extracted histomorphological features from the segmentation maps of H&E-stained WSIs at a single-cell and tissue compartment resolution (e.g. areas, shape metrics, and intensity measurements) [15]. The features were divided into three major histological themes. The first theme was called Neoplastic Nuclear Morphology, capturing metrics on neoplastic nuclei sizes, shapes, nuclear stain intensities, and nuclei-to-chromatin clump proportions. The second theme, Immune Response, focused on quantifying immune cell abundance, localization, and clustering within the biopsies. The third theme, Epithelial Structures, contained information on the structure of epithelial compartments. This theme focused on capturing size, shape, and proportion related features of squamous and glandular epithelial and neoplastic tissue regions, as well as epithelial and neoplastic cell counts.

All features were first computed and aggregated at the slide level using predefined aggregation functions, including the mean and standard deviation of object-level measurements, absolute counts, and proportions relative to tissue areas (Supplementary Table S1). For patients with multiple CIN2 biopsies, slide-level feature values were subsequently summarized to the patient level using fixed aggregation operators, namely the mean, maximum, or sum across all slides belonging to the same patient.

### Clinical and HPV genotyping Features

To complement the histological features, we included five features related to clinical characteristics of the patients or to HPV genotyping results (from now on, clinical/HPV features). Clinical features included colposcopic size of the lesion and entry cytological result that both were previously shown to be predictive of CIN2 regression in this cohort [10]. In addition, we incorporated HPV genotype information derived from Luminex-based genotyping. From the various HPV types detected in our cohort, we selected only two high-risk genotypes with sufficient numbers, HPV16 with 58 (39.7%) patients and HPV31 with 19 (13.0%) patients, for analysis. We also included a categorization based on the number of detected HPV genotypes (0, 1, or >1) per patient as an auxiliary marker. A detailed summary and stratification of these clinical and HPV genotyping features are provided in Table 1.

**Table 1:** The clinical/HPV genotyping characteristics of the 146 women in the study cohort. Clinical and HPV genotype features stratified by outcome groups: All, Quick Regression (Quick Reg), Regression (Reg), and Other (non-regressing or partially-regressing). Key clinical parameters include Entry Pap-Smear cytology results (categorized by severity to Low-grade and High-grade) and Colposcopic Size (categorized by percentage of transformation zone involved). HPV data derived from Luminex-based genotyping includes the overall HPV genotype status (Negative/Positive, including lrHPV and hrHPV breakdown), specific high-risk types HPV16+ and HPV31+, and the Number of HPV Types detected (categorized as 0, 1, or >1). ASC-US, atypical squamous cells of undetermined significance; LSIL, low-grade squamous intraepithelial lesion; ASC-H, atypical squamous cells that cannot exclude high grade squamous intraepithelial lesion; HSIL, high-grade squamous intraepithelial lesion; AGC-NOS, atypical glandular cells not otherwise specified; AGC-FN, atypical glandular cells, favor neoplasia; HPV, human papillomavirus * Single genotypes presented with all women positive for HPV16 or HPV31 categorized as HPV16+ or HPV31+, irrespective of other genotypes detected, multiple infections with other genotypes allowed

| Clinical and HPV genotyping features |  |  |  |  |
| --- | --- | --- | --- | --- |
|  | N (%) |  |  |  |
|  | All | Quick Reg | Reg | Other |
|  | 146 (100%) | 38 (26.0%) | 88 (60.3%) | 58 (39.7%) |
| **Entry Pap-Smear** |  |  |  |  |
| Not Taken | 1 (0.7%) | 0 (0.0%) | 0 (0.0%) | 1 (1.7%) |
| Low-grade cytology | 35 (24.0%) | 11 (28.9) | 20 (22.7%) | 15 (25.9%) |
| - ASC-US | 15 (10.3%) | 5 (13.2%) | 9 (10.2%) | 6 (10.3%) |
| - LSIL | 19 (13.0%) | 6 (15.8%) | 11 (12.5%) | 8 (13.8%) |
| - AGC-NOS | 1 (0.7%) | 0 (0.0%) | 0 (0.0%) | 1 (1.7%) |
| High-grade cytology | 110 (75.3%) | 27 (71.1%) | 68 (77.3) | 42 (72.4%) |
| - ASC-H | 53 (36.3%) | 15 (39.5%) | 37 (42.0%) | 16 (27.6%) |
| - HSIL | 57 (39.0%) | 12 (31.6%) | 31 (35.2%) | 26 (44.8%) |
| **Colposcopic Size** |  |  |  |  |
| 0-3 (<25%) | 67 (45.9%) | 25 (65.8%) | 51 (58.0%) | 16 (27.6%) |
| 4-6 (26%-50%) | 49 (33.6%) | 10 (26.3%) | 28 (31.8%) | 21 (36.2%) |
| 7-12 (>50%) | 30 (20.5%) | 3 (7.9%) | 9 (10.2%) | 21 (36.2%) |
| **HPV Genotype** |  |  |  |  |
| Negative | 20 (13.7%) | 11 (28.9%) | 17 (19.3%) | 3 (5.1%) |
| Positive | 126 (86.3%) | 27 (71.1%) | 71 (80.7%) | 55 (93.2%) |
| - lrHPV+ | 7 (4.8%) | 3 (7.9%) | 6 (6.8%) | 1 (1.7%) |
| - hrHPV+ | 119 (81.5%) | 24 (63.2%) | 65 (73.9%) | 54 (93.1%) |
| - HPV16+ | 58 (39.7%) | 7 (18.4%) | 29 (33.0%) | 30 (51.7%) |
| - HPV31+ | 19 (13.0%) | 9 (23.7%) | 13 (14.8%) | 6 (10.3%) |
| - other hrHPV+ | 61 (41.8%) | 17 (44.7%) | 36 (40.9%) | 25 (43.1%) |
| **Number of HPV Types** |  |  |  |  |
| 0 | 20 (13.7%) | 11 (28.9%) | 17 (19.3%) | 3 (5.1%) |
| 1 | 78 (53.4%) | 20 (52.6%) | 42 (47.7%) | 36 (62.1%) |
| >1 | 48 (32.9%) | 7 (18.4%) | 29 (33.0%) | 19 (32.8%) |

Evaluation of Feature Set Predictive Performance

To evaluate the predictive power of the extracted histological and clinical feature themes, we assessed each theme-specific feature set separately to train machine learning classifiers with the AutoGluon framework for its robust performance on tabular classification tasks. [16]. To obtain a robust estimate of model performance, we used repeated stratified cross-validation. Specifically, a four-fold cross-validation was repeated four times with different random splits, resulting in a total of 16 train–test evaluations for each prediction task (regression and quick regression). Stratified sampling was applied in all splits to preserve the class distribution across folds, which was particularly important given the limited sample size and class imbalance in the dataset.

The predictive performance of the classifiers was comprehensively evaluated with various performance metrics, including Area Under the Receiver Operating Characteristic curve (AUROC), Area Under the Precision-Recall Curve (AUPRC), weighted F1 score, accuracy, balanced accuracy, precision, and recall. This set of metrics was chosen to provide a balanced view of model performance, as single metrics can be misleading in imbalanced datasets and give more weight to performance of the majority class. Models that only predicted the majority class on individual cross-validation folds were discarded as non-predictive. Throughout the text, we report the AUROC as our primary measure of predictive power.

### Feature Importances

We computed importance for every feature in each biological theme with a post-hoc feature importance analysis using SHAP (SHapley Additive exPlanations) values [17]. Following the model evaluation scheme described above, SHAP values were computed on the held-out test splits for each fold, provided the trained classifier was not purely random (i.e., did not exclusively predict the majority class). This repeated cross-validation approach yielded a comprehensive representation of SHAP values, from which the final set of highly predictive features for each biological theme was identified.

### Feature Scoring Heuristic

To obtain a definitive ranking of feature importances, we developed a composite scoring heuristic that integrates information from the SHAP value distributions and the distribution of feature values across the positive and negative sides of the SHAP origin. The heuristic is based on two assumptions: (i) features that exhibit large and consistently strong absolute SHAP magnitudes on either the positive or negative side of the SHAP distribution play a more influential role in the model prediction, and (ii) feature values that separate cleanly between the positive and negative SHAP regions indicate clearer and less mixed predictive effects.

For each feature, we computed both the mean and the total absolute SHAP magnitude separately for the samples with positive SHAP values and for those with negative SHAP values, allowing us to quantify its average and overall effect strength in both directions. Next, we evaluated the distribution of feature values on each side of the SHAP origin by binning the feature values into two categories using Fisher–Jenks optimal classification [18]. This allowed us to quantify how strongly one value category (high or low) dominated within the positive and negative SHAP regions. Features whose value distributions showed minimal mixing between the positive and negative SHAP regions were given higher scores, while greater mixing resulted in reduced scores.

The final feature score was computed as the product of (i) the combined SHAP magnitude term and (ii) the two mixing coefficients representing the purity of feature values on the positive and negative SHAP sides. Features with high predictive contribution and minimal mixing therefore received the highest scores; for example, a feature where high values consistently appear on the positive SHAP side and low values on the negative side would be strongly ranked. A full description of the scoring components is provided in the Supplementary Material.

### Iterative Feature Selection

Feature selection was carried out in multiple stages. First, we trained AutoGluon classifiers separately for each feature theme to evaluate their individual predictive potential. Based on these theme-specific models, SHAP-based scores were computed for all features within each theme, and features were ranked according to the heuristics. For each theme, we then applied the Fisher–Jenks optimal classification to retain only the highest-ranked features.

The selected features from all themes were subsequently used to train a new set of AutoGluon classifiers, providing a combined model that incorporated information across themes. Using SHAP-based scoring again on the combined feature set, we identified the ten highest-ranking features overall, which were then used to train the final set of AutoGluon classifiers.

### Feature Synergies and Redundancies

Many of the selected features encode either complementary (synergistic) or redundant information, likely due to the inherent biological relationships. Following feature selection, we employed the Facet (v2.1.1) library (https://github.com/BCG-X-Official/facet) to compute the Synergy-Redundancy-Independence (S-R-I) decomposition, quantifying feature redundancy and synergy using SHAP values and SHAP dependency interactions [19]. Since SHAP dependency values require tree-based models [20], we trained a set of Random Forest and Extra Trees classifiers using a 4×4 repeated stratified cross-validation procedure. To ensure reliable decomposition results, only classifiers achieving an AUROC ≥ 0.7 were included in the S-R-I analysis, thus guaranteeing that only models with sufficient predictive performance contributed to the evaluation.

### Model Comparison

To benchmark the predictive performance of our interpretable feature-based approach, we compared it against several state-of-the-art Multiple Instance Learning (MIL) deep learning models. We trained large and small variants of CLAM, TransMIL, and AttentionMIL models to predict both overall lesion regression and quick regression outcomes [21–23]. Model training was executed using the Slideflow framework (v3.0.0) [24]. For the underlying feature extraction from the Whole Slide Images (WSIs), we employed two different backbones: UNI and Virchow [25,26]. Every MIL model was trained and validated using the same cross-validation setup utilized for the AutoGluon classifiers, ensuring a direct and unbiased assessment of the MIL models’ predictive accuracy against our interpretable feature-based machine learning pipeline.

## Results

Our aim was to extract histological features from the neoplastic lesion and its microenvironment using diagnostic biopsies of actively surveilled CIN2 lesions and see if these features would predict 1) the regression potential of the lesions within two years and 2) the regression potential of the lesions within one year (quick regression).

### Panoptic Segmentation and Histological Feature Extraction

To enable the extraction of histological features, we first applied panoptic segmentation to H&E-stained diagnostic biopsies using the Histolytics library’s Cellpose model implementation, which was pre-trained on cervical H&E data [13,15].

Using the panoptic segmentation maps of the biopsies, we extracted 76 histological features for each WSI (Supplementary Table S1). The extracted features were designed to represent the key characteristics of the lesions, including tissue morphology and cellular changes indicative of cellular transformation. We hypothesized that these measurements, along with the quantification of microenvironmental responses such as inflammatory nuclei accumulation close to lesion or infiltration into the lesion, could predict the regression potential of CIN2 lesions. The features were categorized based on different histological themes: Nuclear Morphology of Neoplastic Cells (Fig. 1A), Immune Response (Fig. 1B) and Lesion and Epithelial Structure (Fig. 1C).

**Figure 1.**
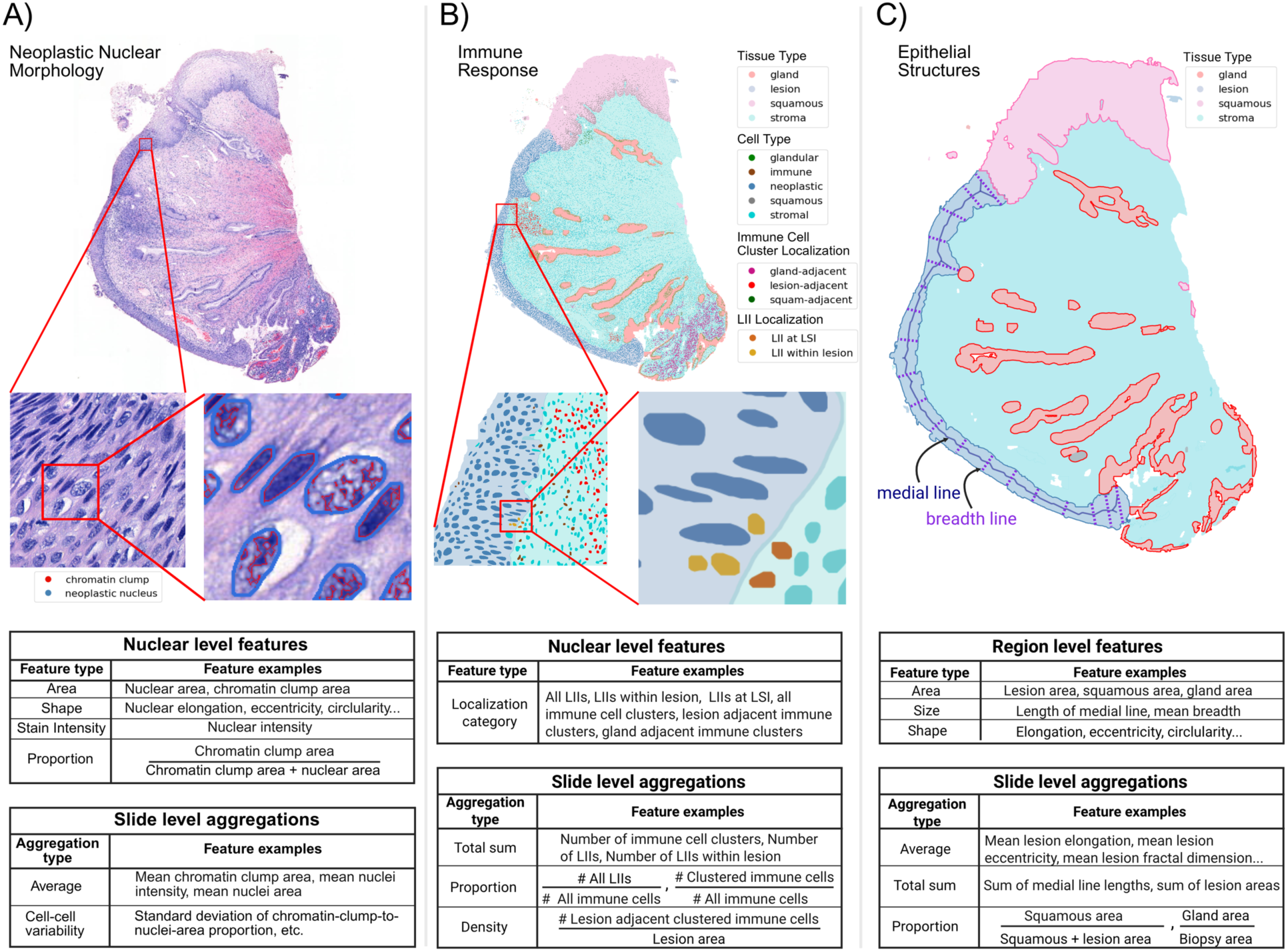
Example illustrations of the extracted histological features by histological theme. Visual illustrations of the three major histological feature themes and slide-level aggregation methods used to quantify high-resolution histological information from Cervical Intraepithelial Neoplasia grade 2 (CIN2) biopsies. **A) Neoplastic Nuclear Morphology Features:** Captures single nuclear level measurements of neoplastic nuclei sizes, shapes, chromatin stain intensities, and nuclei-to-chromatin clump proportions. The nuclear-level features aggregated into slide-level averages and cell-cell variabilities. **B) Immune Response Features:** Focuses on spatial quantification of inflammatory nuclei by categorizing them based on their participation in immune clusters and location within the biopsy with respect to healthy and dysplastic epithelial tissues. These spatial measurements are aggregated to slide-level information via nuclear counts, proportions, density measures, and cluster-level metrics like inflammatory cluster counts. **C) Epithelial Structures Features:** Quantifies the spatial and structural characteristics of healthy and dysplastic epithelial tissue types within the CIN2 biopsies. Measurements include the sizes, areas, and shapes of tissue compartments. These measurements are aggregated to slide-level information via averages, sums, and tissue type proportions between the neoplastic lesion and squamous and glandular epithelial compartments. Created in BioRender. Lehtonen, O. (2026) https://BioRender.com/wqsw5d6 LII, lesion infiltrating immune cells, LSI, lesion stroma interface

### Feature Predictiveness Evaluation and Selection

To assess the predictive contribution of different feature themes, we evaluated histological feature themes and Clinical/HPV features separately using cross-validation (Fig 2A). In this setting, AutoGluon classifiers were trained on the training splits only, and predictive performance was evaluated on the corresponding held-out test splits. Using this approach, the Clinical/HPV feature set showed the strongest predictive performance, with mean AUROC values of 0.78 for regression and 0.74 for quick regression. Out of the histological feature themes, the Epithelial Structures were the most predictive (AUROC reg/quick: 0.64/0.65), followed by Immune Response (0.63/0.64) and Neoplastic Nuclear Morphology (0.63/0.56) as shown in Supplementary Tables S2 and S3.

**Figure 2.**
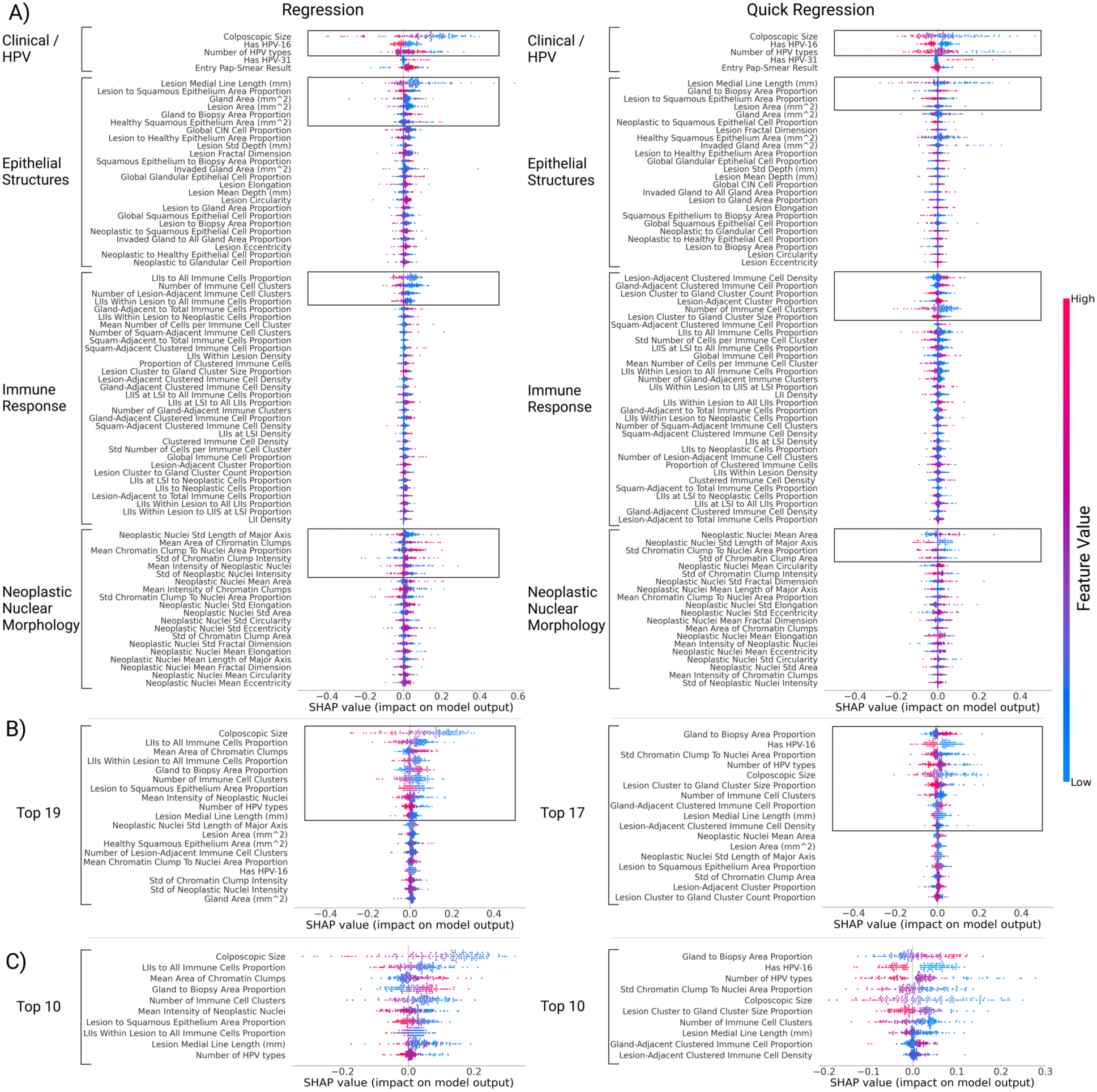
SHAP value-based feature selection process to identify the top 10 most important features for lesion regression and quick regression. The beeswarm plots show the ranked feature importances of the AutoGluon classifiers. Each point in a beeswarm plot represents a single SHAP value of a feature computed from a test set example. The points on the right and left sides of the origin depict how much the feature influenced in classifying a test example to regression/quick regression (right side) or non-regression/non-quick regression class (left side). The individual features are ranked and selected using our SHAP scoring heuristic (selected features highlighted by black boxes). **A)** In the first step, we trained AutoGluon classifiers separately for each of the distinct histological feature theme sets and for the clinical/HPV genotyping features, selecting 19 features and 17 features for regression and quick regression prediction respectively. **B)** Ranked feature importances after the second AutoGluon training and feature selection iteration for both regression and quick regression prediction tasks. **C)** Ranked feature importances of the top 10 features at the final AutoGluon training and feature selection iteration for both regression and quick regression prediction tasks.

We then applied an iterative feature selection procedure to test whether the feature themes’ predictive power is complementary using SHAP values computed from cross-validated models trained within each feature theme. The highest-scoring features from each theme were retained, reducing the feature space from 81 to 19 features for regression prediction and 17 features for quick regression prediction (Fig. 2B).

Using these reduced feature sets, a new set of AutoGluon classifiers was trained from scratch and evaluated using the same repeated stratified cross-validation scheme. Feature importance was then recomputed within this reduced feature space using SHAP values aggregated across cross-validation test folds, and the ten highest-ranking features were selected. Final AutoGluon classifiers were subsequently trained using only these ten features and evaluated using cross-validation with mean AUROC values of 0.82 for regression and 0.79 for quick regression (Fig. 2C, Supplementary Tables S2 and S3; Supplementary Figs. S1 and S2). Because feature selection at each stage was based on importance estimates aggregated across all cross-validation folds rather than being performed independently within each training fold, the resulting performance estimates should be interpreted as mildly optimistic and reflective of model selection performance rather than fully unbiased generalization performance.

To place the combined feature selection results into context, we additionally evaluated the predictive contribution of histological features in isolation by repeating the feature selection and model training procedure after excluding Clinical/HPV variables. Using histological features alone, the models retained measurable prognostic signal for regression (AUROC 0.70), indicating that quantitative morphology extracted from routine H&E slides contains information relevant to lesion outcome. However, predictive performance was consistently inferior to models based on Clinical/HPV features, and model stability across cross-validation splits was lower, particularly for quick regression prediction (Supplementary Figs. S3–S5). These results indicate that histological features provide complementary rather than standalone predictive value in this setting.

We additionally trained deep learning–based MIL models with state-of-the-art foundation backbones using the same cross-validation protocol to provide a reference point for end-to-end whole-slide image prediction in this cohort [25,26]. These models showed limited predictive performance for both regression and quick regression, highlighting the inherent difficulty of predicting CIN2 regression directly from raw WSIs alone (Supplementary Tables S4 and S8). In this setting, the results suggest that modeling explicitly defined histological features together with clinical information offers a viable and interpretable alternative for CIN2 regression prediction.

### Top 10 predictive features for CIN2 regression and quick regression

The resulting top ten clinical/HPV and histological features for prediction of CIN2 regression and quick regression are depicted in Table 1 and 2, respectively. To investigate the complementary nature of the top ten features with each other or whether the features contain redundant information between them, we also quantified the level of pairwise independence, redundancy, and synergy of the features, using SHAP vector decomposition [19].

The most important feature to predict overall regression in two years was the colposcopic size of the lesion, with smaller lesion sizes strongly predicting regression. Of HPV related features, no detected HPV or only one detected type (as opposed to several genotypes) were predictive with no detected HPV types having a strong predictive tendency towards regression. Out of the histological features, three immune response features were also key predictors, with high redundancies with each other. Namely, a low proportion of immune cells within and at the interface of the lesion and low number of immune cell clusters associated with regression. In terms of neoplastic nuclear morphology, larger chromatin clump area and lower mean grayscale intensity, indicating more uniformly distributed chromatin and darker nuclear staining, were predictive of overall regression. In line with small colposcopic size of the lesion, out of the features describing the composition of the epithelial compartments, the short length of the medial line of the lesion and low lesion to healthy squamous epithelium–ratio, both indications of small histological size of the lesion, were predictive. Also, a high proportion of glandular structures within the biopsy predicted regression, with redundancies with immune response features. Interesting synergistic effects linking the number of HPV types and colposcopic size together was detected, as well as a synergistic effect between the lesion to squamous epithelium proportion and mean grayscale intensity of neoplastic nuclei (Table 2).

**Table 2:** Top 10 features for predicting CIN2 regression in two years in order of importance. The “Redundancy” column lists features that share more than 10% overlapping predictive information with others, and the “Synergy” column maps the degree to which the model combines information from one feature to another. The clinical/HPV features are highlighted in bold. Colposcopic size and the mean area of chromatin clumps appeared as independent predictors, whereas several immune-related and tissue-structure features showed measurable redundancy with one another. Notably, a subset of features demonstrated synergistic effects, including interactions between nuclear intensity and lesion-to-squamous proportions and between HPV genotype diversity and colposcopic size, indicating that certain feature combinations provided additional predictive value beyond their individual contributions. LII, lesion infiltrating immune cells

| Top 10 Features for Prediction of CIN2 Regression in two years |  |  |  |  |  |
| --- | --- | --- | --- | --- | --- |
| Rank | Feature | Description | Predicted Regression | Redundancy (>10%) | Synergy (>5%) |
| 1 | **Colposcopic Size** | Proportion of involved transformation zone | Low values |  |  |
| 2 | LIIs to All Immune Cells Proportion | Proportion of immune cells within 64 μm of neoplastic cells relative to all immune cells | Low values | LIIs Within Lesion to All LIIs Proportion (62.4%), Gland to Biopsy Area Proportion (10.8%) | |
| 3 | Mean Area of Chromatin Clumps | Mean area of chromatin clumps within the nuclei of neoplastic nuclei | High values |  |  |
| 4 | Gland to Biopsy Area Proportion | The proportion of glandular area relative to the total biopsy area | High values | LIIs to All Immune Cells Proportion (10.8%), LIIs Within Lesion to All LIIs Proportion (17.0%) |  |
| 5 | Number of Immune Cell Clusters | The number of immune cell clusters in the biopsy | Low values | Lesion Medial Line Length (32.6%) |  |
| 6 | Mean Intensity of Neoplastic Nuclei | Mean grayscale intensity of neoplastic nuclei | Low values |  | Lesion to Squamous Epithelium Area Proportion (8.1%) |
| 7 | Lesion to Squamous Epithelium Area Proportion | The proportion of lesion area relative to healthy squamous epithelium area. | Low values |  |  |
| 8 | LIIs Within Lesion to All LIIs Proportion | The proportion of immune cells within the lesion relative to all immune cells. | Low values | LIIs to All Immune Cells Proportion (62.2%), Gland to Biopsy Area Proportion (16.9%) |  |
| 9 | Lesion Medial Line Length | The length of the medial line of the lesion. | Low values | Number of Immune Cell Clusters (32.7%) |  |
| 10 | **Number of HPV types** | The number of unique HPV types detected, categorized as 0 (none), 1 (one type), or >1 (multiple types) | Low values |  | Colposcopic size (6.3%) |

Somewhat surprisingly, the strongest predictor of quick regression was a high proportion of glandular tissue within the biopsy, possibly mirroring the localization of the lesion in the transformation zone. The second and third most important features were the absence of an HPV16 infection and none or only one detected HPV genotype. While in the prediction of overall regression the LIIs and number of immune cell clusters were important, in the prediction of quick regression the important immune response features were related to the spatial localization, number and size of immune cell clusters (with high redundancies between the features related to localization of the clusters). Feature synergies were seen between histological features and the HPV genotype related features. Especially HPV16 status shared synergies with features from all histological themes outlining the synergy between histological and etiological features (Table 3).

**Table 3:** Top 10 features for predicting CIN2 quick regression. The “Redundancy” column lists features that share more than 10% overlapping predictive information with others, and the “Synergy” column maps the degree to which the model combines information from one feature to another. The clinical/HPV features are highlighted in bold. The most influential predictors included gland-to-biopsy area proportion and HPV-related variables, with HPV-16 status and the number of HPV types acting as independent contributors. Immune-cluster–related features showed extensive redundancy with one another, particularly among gland-adjacent and lesion-adjacent cluster characteristics. Synergistic effects were observed between chromatin-clump area variability and HPV-16 status, lesion medial line length and HPV-16 status, and lesion-adjacent immune cluster density with HPV-related variables, indicating that specific combinations of histological and clinical variables contributed additional predictive value beyond their individual effects. LII, lesion infiltrating immune cells

| Top 10 Features for Prediction of CIN2 Quick Regression |  |  |  |  |  |
| --- | --- | --- | --- | --- | --- |
| Rank | Feature | Description | Predicted Quick Regression | Redundancy (>10%) | Synergy (>5%) |
| 1 | Gland to Biopsy Area Proportion | The proportion of glandular area relative to the total biopsy area | High values | Lesion-Adjacent to Gland-Adjacent Immune Cluster Size Proportion (10.6%), Gland-Adjacent Clustered Immune Cell Proportion (11.0%) |  |
| 2 | **HPV-16 status** | HPV16 detected in genotyping | Negative HPV-16 status |  |  |
| 3 | **Number of HPV types** | The number of unique HPV types detected in genotyping, categorized as 0 (none), 1 (one type), or >1 (multiple types) | None or One detected HPV type |  |  |
| 4 | Standard Deviation of Chromatin Clump to Nuclei Area Proportion | The cell-to-cell variability in chromatin-clump-to-nuclei area proportion | Low values |  | HPV-16 status (6.3%) |
| 5 | **Colposcopic size** | Proportion of involved transformation zone | Low values |  |  |
| 6 | Lesion-Adjacent to Gland-Adjacent Immune Cluster Size Proportion | The size (number of cells) of immune cell clusters located near the lesion relative to those located near glands | Low values | Gland to Biopsy Area Proportion (10.6%), Number of Immune Cell Clusters (10.0%), Gland-Adjacent Clustered Immune Cell Proportion (86.2%), Lesion-Adjacent Clustered Immune Cell Density (11.3%) |  |
| 7 | Number of Immune Cell Clusters | The number of immune cell clusters in the biopsy | Low values | Lesion-Adjacent to Gland-Adjacent Immune Cluster Size Proportion (10.0%), Lesion Medial Line Length (21.9%) |  |
| 8 | Lesion Medial Line Length | The length (mm) of medial line of the lesion | Low values | Number of immune cell clusters (21.9%) | HPV-16 status (7.5%) |
| 9 | Gland-Adjacent Clustered Immune Cell Proportion | The proportion of gland-adjacent clustered immune cells relative to all clustered immune cells | High values | Gland to Biopsy Area Proportion (11.6%), Lesion-Adjacent to Gland-Adjacent Immune Cluster Size Proportion (85.1%) |  |
| 10 | Lesion-Adjacent Clustered Immune Cell Density | The number of lesion-adjacent clustered immune cells relative to stromal area | Low values | Lesion-Adjacent Cluster to Gland-Adjacent Cluster Size Proportion (10.8%) | HPV-16 status (10.5%), Number of HPV types (10.0%) |

## Discussion

H&E-stained slides of histological biopsies are the cornerstone of cervical cancer precursor lesion diagnostics and widely available for CIN2 lesions and patients, but pathologists are currently not able to predict the regression potential of the lesions from the neoplastic or tumor microenvironment related features visible in them. We extracted 76 interpretable histological features from diagnostic biopsies of women under active surveillance for CIN2 and complemented these with five clinical and HPV-related features, resulting in a total of 81 features. These features were then used within a ML framework to identify the most informative histological and clinical variables for predicting regression during two-year follow-up (regression) and regression within one year (quick regression). Clinical and HPV genotyping related variables alone showed good predictive power for both overall and quick regression of CIN2 lesions. Still, the highest performance in regression prediction was achieved when combining the most informative features from both histological and clinical/HPV-related categories.

To our knowledge, this is the first study to use histological features in routine diagnostic H&E samples alone and in combination with HPV genotyping data and clinical features to predict the regression potential of CIN2 lesions. As reported earlier for this cohort and in line with previous studies, small colposcopic lesion size (<50 % of the transformation zone) was highly predictive of regression potential (1^st^ in overall regression, 5^th^ in quick regression) [10,27]. Further, no or only one detected HPV type detected in extended genotyping, and in the histology of the biopsies, short medial length of the lesion (small histological lesion size), low number of immune cell clusters, and high proportions of glandular tissues predicted both regression and quick regression. Also features related to nuclear morphology were important for both overall and quick regression.

Pathologists use nuclear abnormalities (hyperchromasia, coarse chromatin, irregular membranes), an increased nucleus:cytoplasm ratio as diagnostic characteristics of high-grade CIN lesions, but they are not known to hold predictive power in CIN2 lesions. In our study, the key nuclear characteristics were linked not to the size or shape of the neoplastic nuclei, but to the pattern of chromatin distribution, reflecting the underlying structure of the chromatin. Previous tissue-based studies have found that immunohistochemical biomarkers indicative for a productive HPV infection (HPV E4 protein expression) hold some predictive potential in high-grade CIN lesions whereas the commonly used markers of transforming HPV infection (p16INK4a and Ki-67 expression) have conflicting results [28–31]. Our results indicate that the structural changes in the chromatin occurring in transforming HPV infections might also be detectable solely based on H&E histology, without molecular diagnostic tools. Moreover, investigating these neoplastic nuclei further with spatial sequencing methods could provide valuable insights into underlying transcriptional events.

Previous biopsy-based studies have also found that ratios of different immune cells phenotypes predict regression potential of CIN2 lesions [32]. Despite the limitations of our model to distinguish between different immune cell types, predictive immune response features were found in both our analyses. Low numbers of immune cell clusters were predictive in both time frames, low proportion of LIIs predicted overall regression, and low density and small size of immune cell clusters close to the lesion, but high proportions of immune cell clusters close to endocervical glands in the biopsies predicted quick regression. These results highlight the importance of the spatial patterns of immune cell distribution.

The absence of HPV16 was 2^nd^ most important feature in quick regression prediction and shared synergies with features from all histological themes. Also, in the prediction of overall regression in two years, the number of detected HPV types was especially synergistic with histological features from all inspected themes. These findings highlight the importance of a holistic view between etiological factors (HPV) and histological features, mirroring the effects of HPV in neoplastic cells/lesion and the response they elicit (or not) in the microenvironment. Moreover, as we show, even state-of-the-art H&E-based DL-models fell short of achieving good predictive accuracy, highlighting that a holistic, cross-feature integration is essential in achieving a good predictive power when data is constrained and noisy.

While our findings demonstrate promising predictive potential, the reported performance should be interpreted with caution due to potential selection-induced bias. Although feature importance was estimated using cross-validation and models were retrained from scratch at each stage, feature selection was based on importance scores aggregated across cross-validation folds rather than being performed independently within each training split. Consequently, the final AUROC values reflect model selection performance and may be mildly optimistic with respect to true generalization. Future studies using fully nested feature selection or independent validation cohorts will be required to obtain unbiased estimates of predictive accuracy. In addition, the cohort size was moderate, which likely contributes to the relatively modest predictive performance observed for individual histological feature themes and may limit the generalizability of the identified feature associations. Accordingly, external validation in larger, multi-center datasets will be required to assess the robustness and clinical applicability of the proposed approach. Although all slides underwent stain normalization and visual quality control, residual variability in staining intensity and section quality may still influence quantitative feature extraction.

In conclusion, we show that the routine H&E histology of diagnostic biopsies contains relevant information that predicts the natural history of CIN2 lesions. Although the predictive strength of histology alone was lower compared to clinical and HPV features, they provide complementary information that enhances prediction performance when integrated into combined analyses. Compared to molecular analysis, H&E histology is routinely widely available and is translatable to features that could be used in algorithms guiding treatment decisions. Further, these features provide valuable cues to future tissue-based studies on the natural history of CIN2 lesions.

## Data & Code Availability

The datasets generated and analyzed during the study originate from the Helsinki University Hospital (HUS) patient cohort and contain sensitive human subject information. Due to institutional policy and patient confidentiality restrictions, these data are not publicly available. Access to the data may be requested through formal collaboration with the corresponding authors, subject to institutional ethical approval and data use agreements. The source code used for data processing, feature extraction, model training, and analysis is publicly available on GitHub at https://github.com/okunator/CIN2_regression.

## Acknowledgements

We acknowledge CSC-IT Center for Science, Finland, for computational resources. We also acknowledge the support from Finnish Medical Foundation (recipient I.K) and O.L acknowledges Finnish Society for Colposcopy r.y and the Biomedicum Helsinki Foundation for personal research grants. Additionally, we acknowledge Helsinki Biobank for scanning the whole-slide data used in this work.

## Ethics Approval and Consent to Participate

The study was approved by Helsinki-Uusimaa Hospital District Ethical Committee (131/13/03/03/2013;24/4/2013) on April 24, 2013. All women gave written informed consent.

