## Supplementary Material for "Integrating Quantitative Histology with Clinical Data Improves Prediction of Cervical Intraepithelial Neoplasia Regression"

### Histological Feature Extraction

#### Neoplastic Nuclear Morphology

Neoplastic nuclear morphologies were extracted for high-grade squamous intraepithelial lesion (HSIL) cells, as well as for low-grade squamous intraepithelial lesion (LSIL) cells that may be present in CIN2 biopsies. The extracted nuclear shape metrics included nuclei area, circularity, elongation, compactness, eccentricity, fractal dimension (boundary complexity), convexity, solidity, major axis length, and minor axis length. All features were computed using the Histolytics library [1]. Chromatin-related features were extracted using the chromatin_feats function. Grayscale intensity-based features were computed using the grayscale_intensity_feats function. The rationale for restricting feature extraction to neoplastic cells was that CIN2 represents a non-invasive lesion, and thus, nuclear morphologies in the surrounding microenvironment are less likely to be biologically relevant or interpretable for regression prediction.

#### Epithelial Structures

For epithelial structures, we extracted lesion tissue morphology features using the shape_metric function from the Histolytics library. The extracted features included area, elongation, eccentricity, circularity, major axis length, and fractal dimension. In addition, we computed the lesion medial line length and mean breadth by averaging the lengths of lines perpendicular to the medial line within the lesion. These metrics were obtained using the medial_lines and perpendicular_lines functions from Histolytics. Tissue regions smaller than approximately 160 $\mu m^2$ were excluded prior to feature computation.

#### Immune Response

To quantify immune infiltration, the biopsy tissue was divided into two compartments: the lesion and the lesion–stroma interface (LSI), defined as a 250 µm buffer extending from the lesion into the stroma. Lesion-infiltrating immune cells (LIIs) were classified based on their location within either the lesion or the LSI compartment.

Immune cell clustering was performed using the density_clustering function from the Histolytics library, which applies a DBSCAN-based clustering algorithm with epsilon equal to 200 pixels (approximately 100 µm) and a minimum of 25 cells per cluster. Clusters were further categorized by their localization relative to nearby epithelial structures. Clusters were labeled as adjacent-to-lesion, adjacent-to-gland, or adjacent-to-squamous epithelium if more than 15% of their cells intersected with the corresponding epithelial–stroma interface. The interface region was defined as a 500 µm buffer from the epithelial boundary into the stroma. If a cluster overlapped multiple interfaces, the category was assigned in the order of priority: lesion, squamous, gland. This enabled quantification of cluster features by their histological localization within the biopsy.

### Panoptic Segmentation Inference

Whole-slide inference was performed directly using the Histolytics library, which supports WSI-level panoptic segmentation. Each WSI was divided into non-overlapping 1024 × 1024 pixel tiles at 0.5 MPP for inference. Following inference, the resulting segmentation maps were merged into complete WSI-level outputs. The quality of each segmentation map was reviewed visually using QuPath (v0.5.0), and clear segmentation errors such as misclassification of healthy basal squamous tissue as neoplastic lesion were corrected manually [2].

### AutoGluon Training Configuration

AutoGluon was used to train tabular classifiers for the regression and quick-regression prediction tasks [3]. For each training dataset split, the features and outcome labels were provided to an AutoGluon TabularPredictor object configured to optimize the weighted F1-score. Models were trained using the "best_quality" preset, which enables an extensive model search and stacking of multiple base learners. Automated stacking was activated to improve predictive performance, and decision-threshold calibration was applied during training to optimize classification boundaries. A maximum training time of 10 minutes per model was allowed, while inference time was restricted to a small proportion ("infer_limit" preset 0.005) of the training duration to ensure efficient evaluation.

### SHAP Value Computation

SHAP values were computed using KernelExplainer implemented in the shap python library [4].

### SHAP Value Scoring Heuristic

To rank features for selection, we defined a composite SHAP-based scoring function that combines the magnitude of SHAP values with a measure of how cleanly feature values separate across the SHAP origin.

Let $f$ denote a feature, and let $\varphi_i(f)$ be the SHAP value of feature $f$ for sample $i$ . Define the sets of samples with positive and negative SHAP values for that feature as

$$P\{i \mid \varphi_i(f) > 0\}, N_x = \{i \mid \varphi_i(f) < 0\}$$

**Absolute SHAP contribution term**

For each feature $f$ , we first quantify the SHAP magnitude separately on the positive and negative sides of the SHAP origin. The total absolute SHAP contribution and the mean absolute SHAP contribution are defined as

$$S_+(f) = \sum_{i\in P_f} \mid \varphi_i(f) \mid \ , S_-(f) = \sum_{i\in N_f} \mid \varphi_i(f) \mid$$

$$\mu_+(f) = \frac{1}{P_f}\sum_{i\in P_f} \mid \varphi_i(f) \mid \ , \mu_-(f) = \frac{1}{N_f}\sum_{i\in N_f} \mid \varphi_i(f) \mid$$

The absolute contribution term is then defined as

$$C_{abs}(f) = \mu_+(f)S_+(f) \ + \ \mu_-(f)S_-(f),$$

which favors features that have both large total and large average SHAP magnitudes on either side of the origin.

**Mixing coefficients for positive and negative SHAP regions**

Let $x_i(f)$ denote the value of feature $f$ for sample $i$ . Before computing mixing, feature values are truncated to the 5th–95th percentile range and rescaled (denote the transformed values by $\overline{x_i}(f)$.

To characterize how high and low feature values are distributed across the SHAP origin, we apply Fisher–Jenks classification with two classes, which assigns each sample to one of two bins $b_i(f) \in \{0,\ 1\}$ (interpretable as “low” and “high” values).

For the positive SHAP side, we define the empirical bin proportions

$$p_k^{(+)}(f) = \frac{\left|\{i \in P_f | b_i(f) = k\}\right|}{|P_f|}, k \in \{0,1\}$$

and analogously for the negative SHAP side

$$p_k^{(-)}(f) = \frac{\left|\{i \in N_f | b_i(f) = k\}\right|}{|N_f|}, k \in \{0,1\}$$

Let

$$r_+(f) = \max_{k\in\{0,1\}}^{p_k^{(+)}}(f), r_-(f) = \max_{k\in\{0,1\}}^{p_k^{(-)}}(f)$$

denote the dominant-bin proportions on the positive and negative sides, respectively.

The mixing coefficient for the positive side $M_+(f)$ and the negative side $M_-(f)$ was then determined using the following rules:

1. **Perfect dominance:**
   If all samples on one side of the SHAP origin were assigned to the same bin (that is, no mixing at all), the mixing coefficient for that side was set to a predefined boosting factor $\alpha = 2$ . This reflects the strongest possible separation of feature values.
2. **Strong dominance:**
   If both bins were present but one bin accounted for a large majority of samples, exceeding a predefined dominance threshold $\tau = 0.85$ , the mixing coefficient was computed as the dominant-bin proportion multiplied by the boosting factor $\alpha = 2$ . This gives an intermediate boost for features showing clear but not perfect separation.
3. **Weak or no dominance:**
   If both bins were present and neither bin exceeded the dominance threshold, the mixing coefficient was set equal to the dominant-bin proportion itself. In this case, no boost was applied, reflecting substantial mixing of feature values across the SHAP decision boundary.

**Final feature score**

The final feature score is computed as

$$score(f) = M_+(f)M_-(f)C_{abs}(f)$$

Features with large absolute SHAP contributions and low mixing of feature values on both sides of the SHAP origin therefore receive the highest scores. For example, a feature for which high values consistently appear on the positive SHAP side and low values on the negative SHAP side will tend to have strong separation and a high overall score.

### MIL Model Training

The Slideflow Python library was used to train four MIL architectures with UNI and Virchow backbone feature extractors [5–7]. The architectures were CLAM-big, CLAM-small, AttentionMIL, and TransMIL [8–10]. Each model was trained using a 4×4 repeated stratified cross-validation setup identical to that used for the AutoGluon classifiers. Training ran for 40 epochs with a learning rate of 0.0001, weight decay of 1e–5, batch size of 64, and a one-cycle learning rate scheduler. AUROC was used as the training monitor, and the best model weights were saved automatically. Instance-level loss was computed with cross-entropy, bag-level loss with weighted cross-entropy, and optimization used the Adam optimizer.

### S-R-I Decomposition

To quantify feature–feature interactions, we applied the Facet library (https://github.com/BCG-X-Official/facet) inspection tools to an ensemble of tuned tree-based models. For each prediction task, the selected feature set was used to train RandomForest and ExtraTrees classifiers (sklearn implementations [11]), with hyperparameters optimized using RandomizedSearchCV inside a repeated stratified 4-fold cross-validation scheme (4 repeats). This procedure was run across multiple random seeds, and the best model from each run was retained, yielding an ensemble of 16 fitted classifiers.

For each model in the ensemble, Facet library LearnerInspector was used to compute pairwise redundancy and synergy matrices through the feature_redundancy_matrix and feature_synergy_matrix functions. After all runs were completed, redundancy and synergy values were aggregated by taking the elementwise mean across the all the synergy and redundancy matrices, producing final interaction matrices that reflect stable S–R–I estimates rather than model-specific fluctuations.

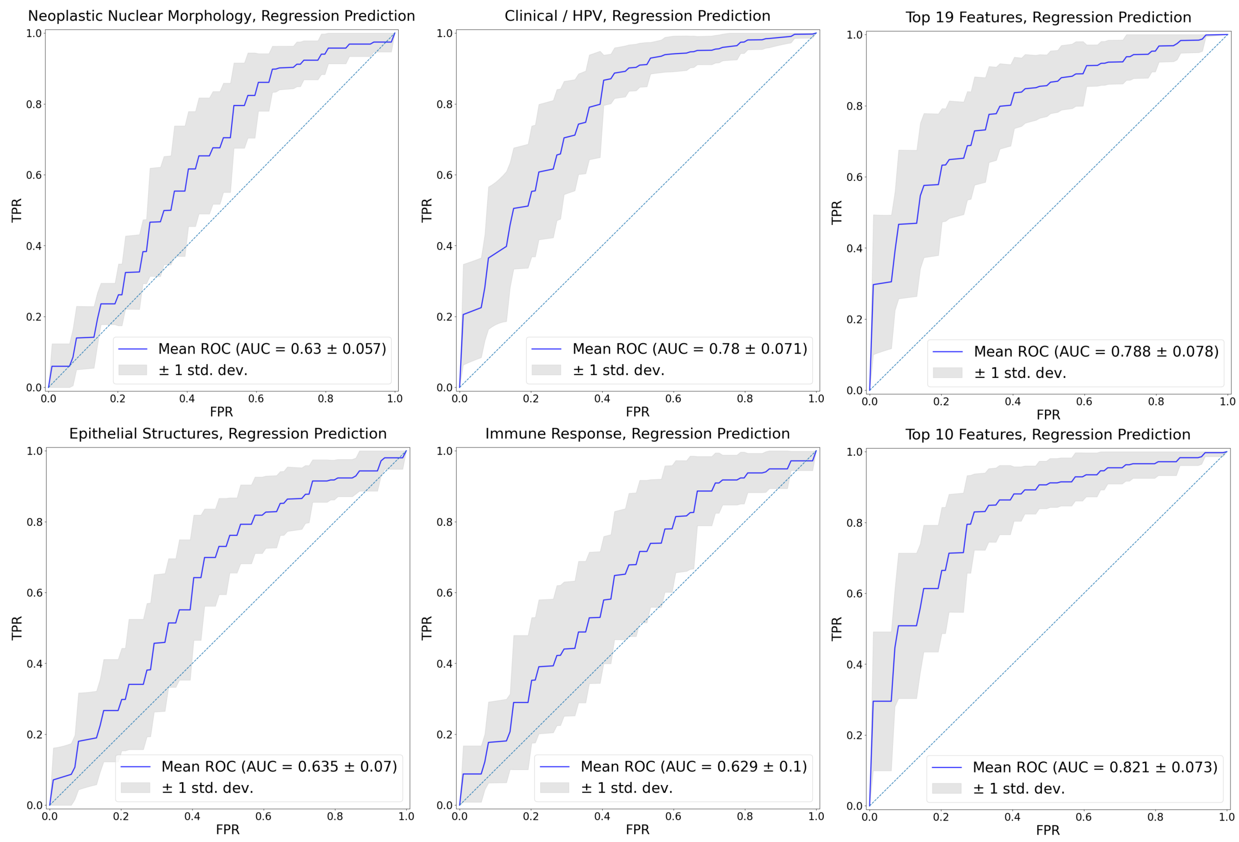


**Supplementary Figure S1: Mean AUROC curves of the Autogluon classifiers for prediction of regression within 2 years.** Histological feature themes i.e. Neoplastic Nuclear Morphology, Epithelial Structures, Immune Response alone obtain only a limited predictive power regression prediction. The Clinical / HPV data, on the other hand, obtains a moderate predictive power alone. After selecting and combining features from the histological feature themes and Clinical/HPV features, we see first a minor ~1% increase in AUROC but when further pruning the features down to top 10, a considerable 4% increase is detected.

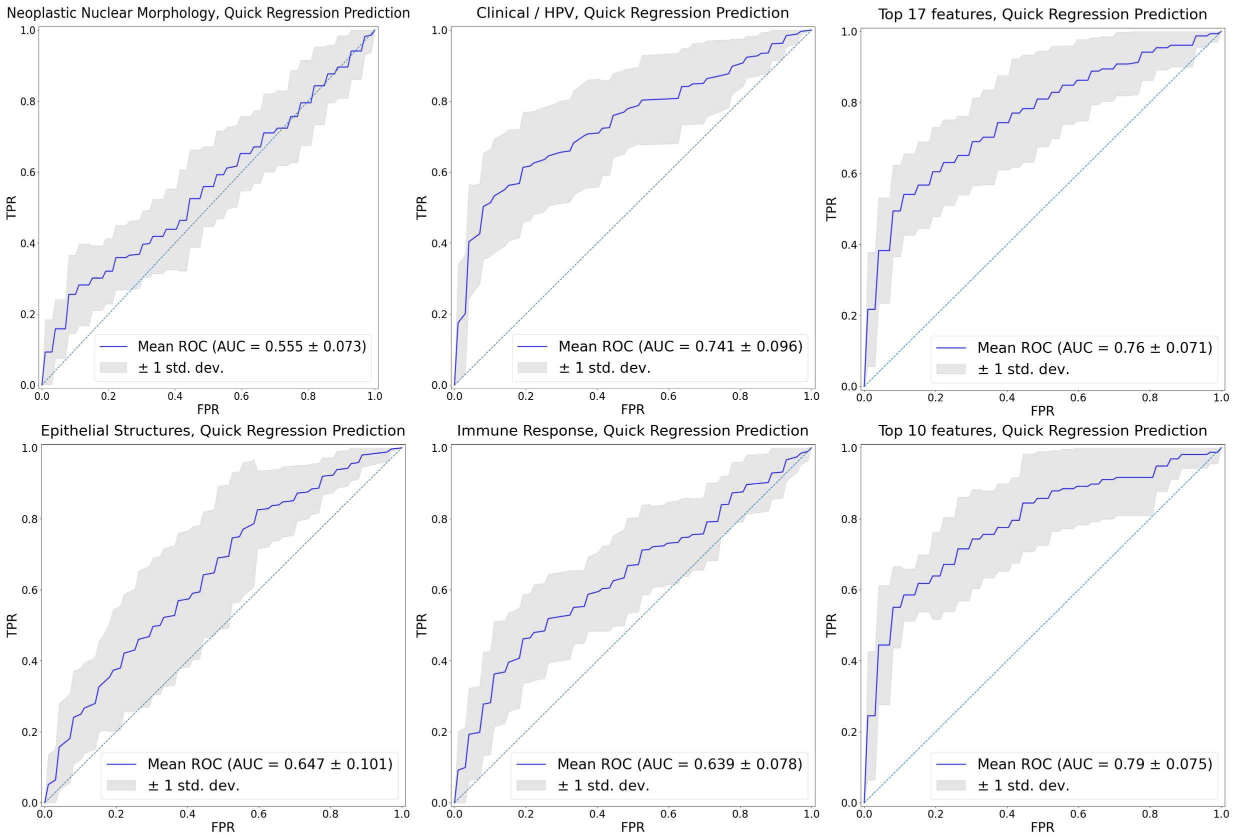


**Supplementary Figure S2: Mean AUROC curves of the Autogluon classifiers for prediction of regression within one year (quick regression)**. Histological feature themes i.e. Neoplastic Nuclear Morphology, Epithelial Structures, Immune Response alone obtain only a limited predictive power in quick regression prediction. The Clinical / HPV data, on the other hand, obtains a moderate predictive power alone. After selecting and combining features from the histological feature themes and Clinical/HPV features, we see first a minor ~2% increase in AUROC but when further pruning the features down to top 10, a considerable 5% increase is detected.

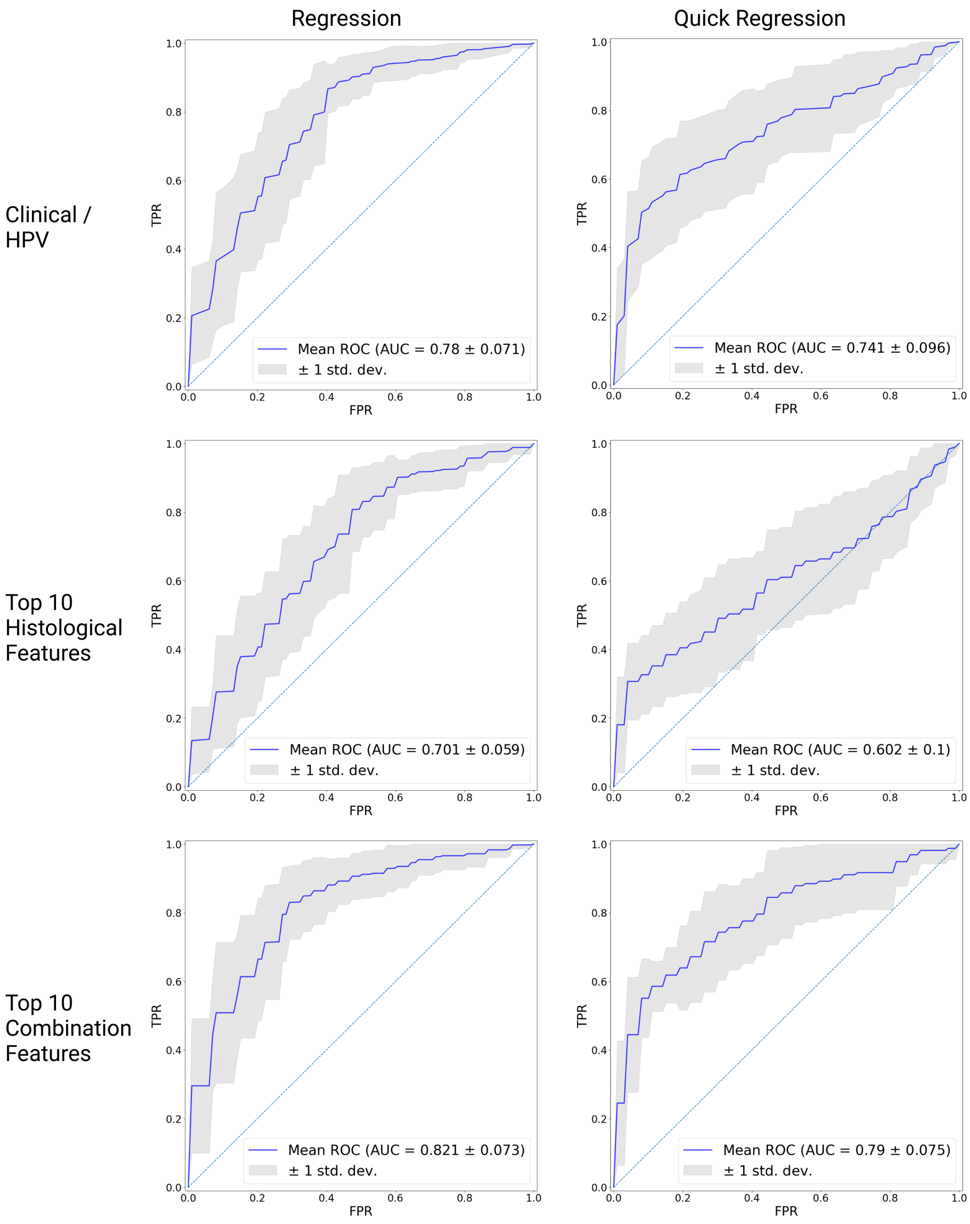


**Supplementary Figure S3: Comparison of mean AUROC curves of Autogluon classifiers for the prediction of regression and quick regression between clinical/HPV genotyping features alone, top 10 histological features alone, and top 10 features overall (clinical/HPV genotyping and histological features in the same model).** Clinical features are stronger predictors alone than histological features. However, histological features alone do carry moderate predictive power in regression prediction, whereas in quick regression prediction, the predictive power of histological features is suboptimal. Top predictive performance is achieved when selecting the best features from both histological and clinical feature categories.

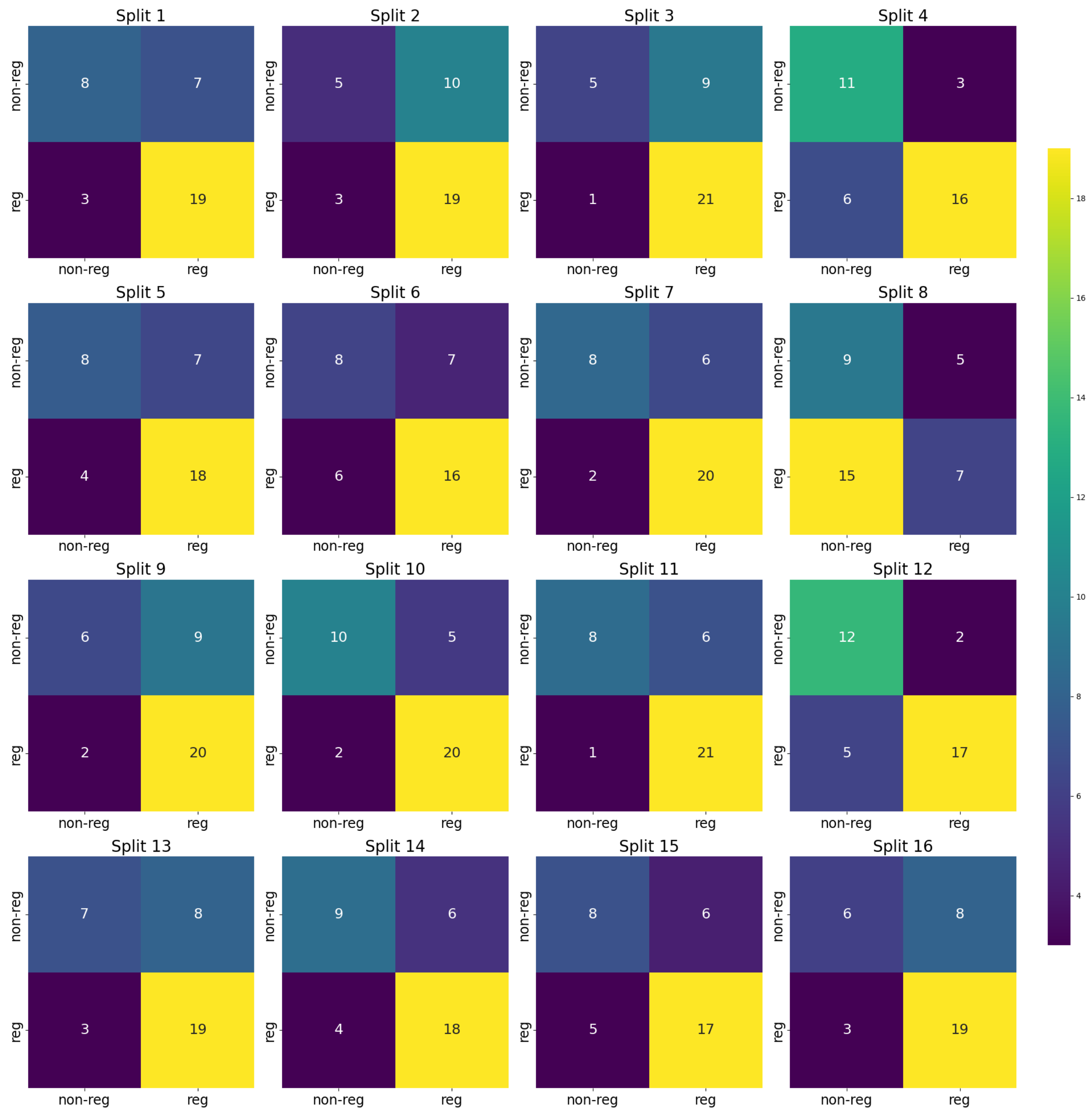


**Supplementary Figure S4: Confusion matrices of Autogluon classifiers for each test split in regression prediction task using only top 10 histological features.** The majority class (regressing patients) is moderately predicted in each test split whereas there is more variation in predicting the minority class (non-regressing patients). In split 8, the classifier learned to mostly predict the minority class whereas in splits 2, 3, 9, 13 and 16 the classifiers learned to mostly predict the majority class.

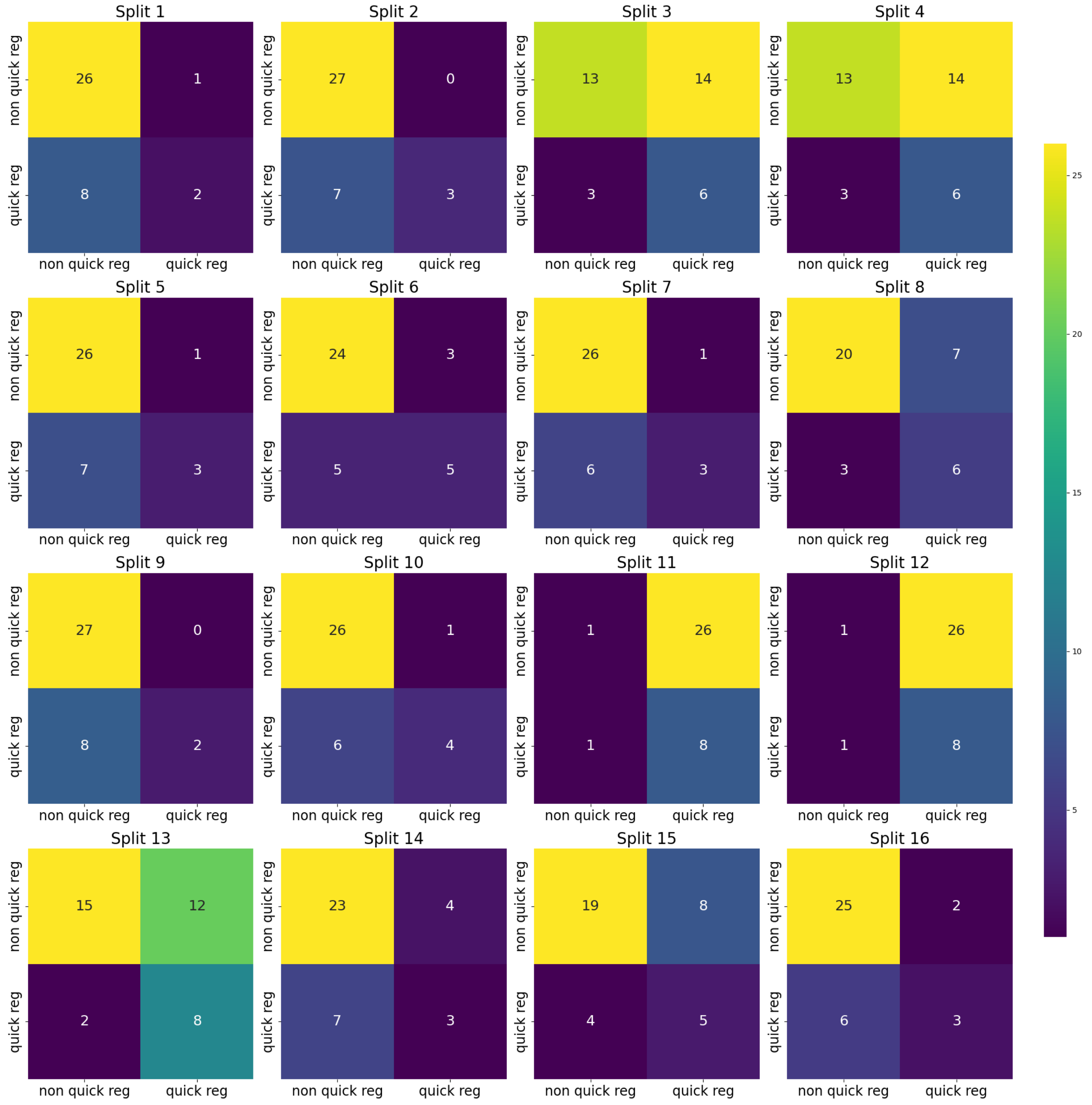


**Supplementary Figure S5: Confusion matrices of Autogluon classifiers for each test split in quick regression prediction task using only top 10 histological features.** The majority class (non-quick regressing patients) is moderately predicted except in test splits 11 and 12 where the classifiers have only learned to predict the minority class (quick regressing patients). In Splits 3, 4, 13, 14, and 15 the classifiers learn to predict well the minority class, although this comes at the expense of precision, leading to many false positives.

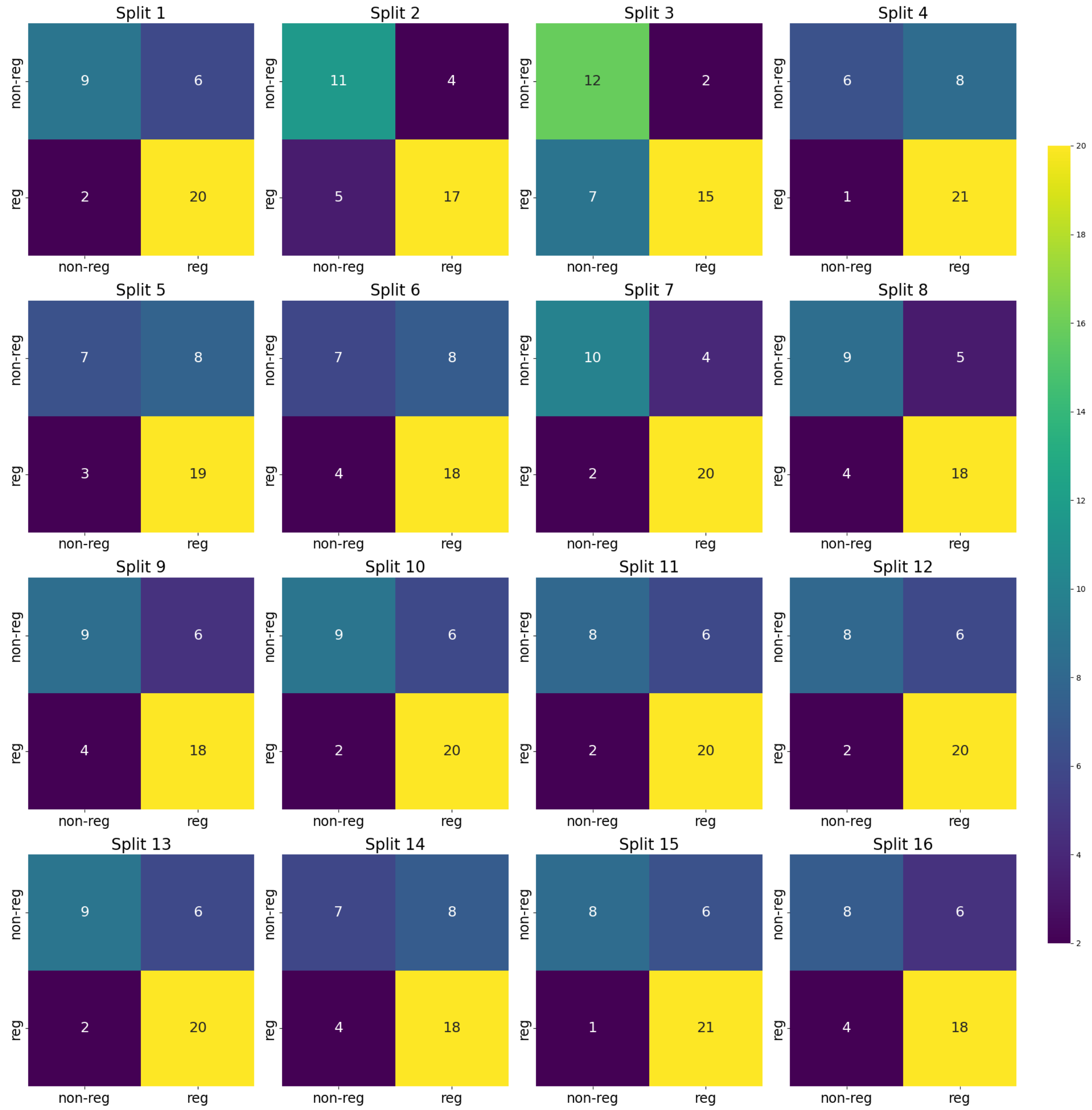


**Supplementary Figure S6: Confusion matrices of Autogluon classifiers for each test split in regression prediction task using only clinical and HPV features.** The majority class (non-regressing patients) is moderately predicted except in test split 3. In Splits 4, 5, 6, and 14 the classifiers learn to predict mostly the majority class.

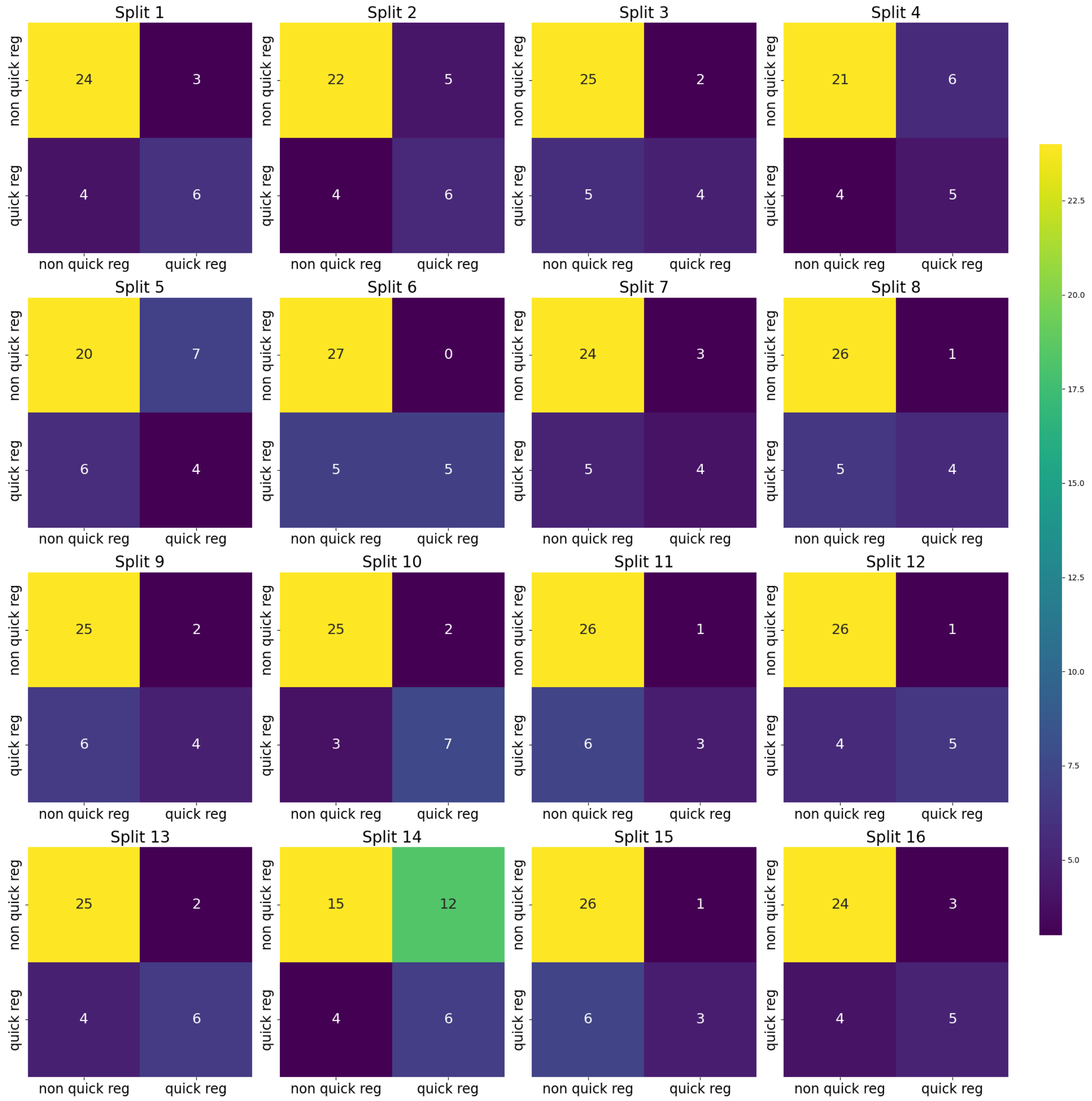


**Supplementary Figure S7: Confusion matrices of Autogluon classifiers for each test split in quick regression prediction task using only clinical and HPV features.** The majority class (non-quick regressing patients) is moderately predicted except in test split 14. For the minority class (quick-regression) in 9 out of 16 splits more than half of the examples were predicted correctly.

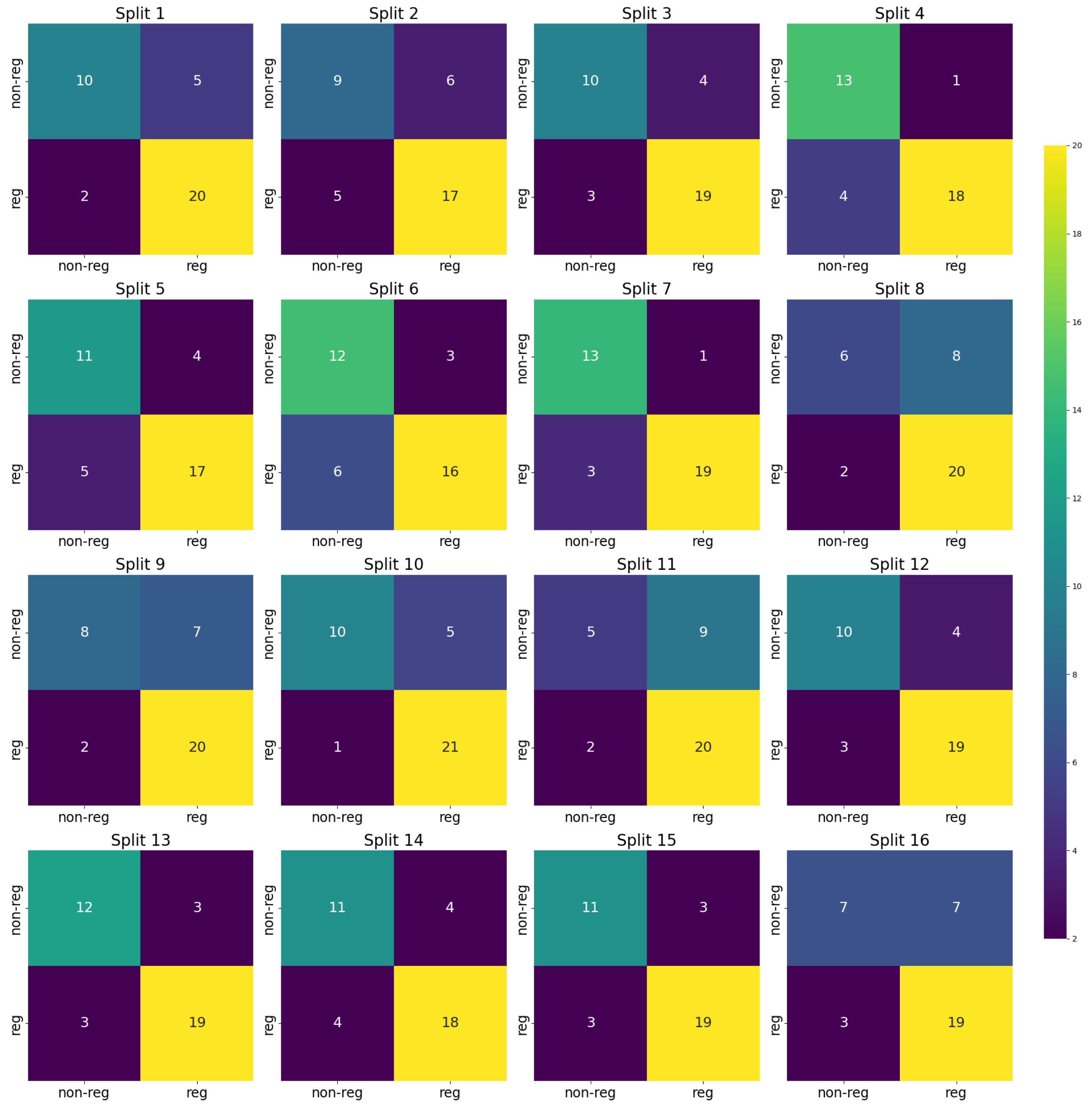


**Supplementary Figure S8: Confusion matrices of Autogluon classifiers for each test split in regression prediction task using the top 10 features of all features (clinical/HPV genotyping and histological features).** For most of the test splits, the classifiers learn a moderate separation of the minority (non-regressing patients) and majority classes (regressing patients). However, in splits 8 and 11 the classifiers learn to predict mostly the majority class.

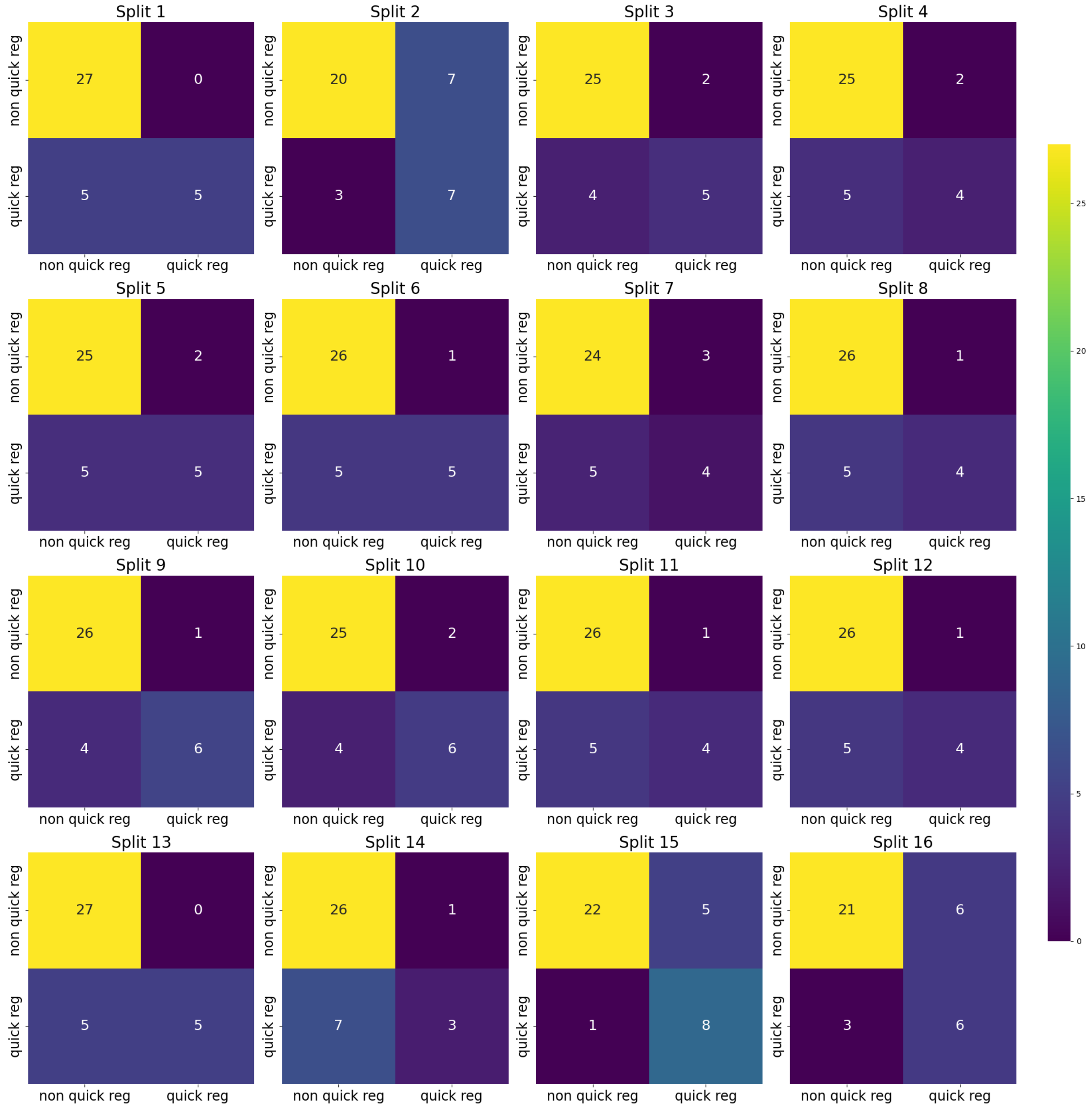


**Supplementary Figure S9: Confusion matrices of Autogluon classifiers for each test split in quick regression prediction task using the top 10 features of all features (clinical/HPV genotyping and histological features).** For most of the test splits, the classifiers learn a moderate separation of the minority (quick regressing patients) and majority classes (non-quick regressing patients). However, in splits 2, 4, 11, and 14 the classifiers learn to predict mostly the majority class.

**Supplementary Table S1: Overview of all extracted histological features.** The table lists each feature together with its corresponding histological theme, slide-level aggregation method, functional description, and a brief interpretation. These features constitute the full set of quantitative predictors derived from the panoptic segmentation maps/WSIs and were used as inputs for feature ranking and model development.

| Feature Set | F1 Weighted | Accuracy | Balanced Accuracy | AUROC | Precision | Recall | AUPRC | AUPRC Above Baseline |
|---|---|---|---|---|---|---|---|---|
| Immune Response Features | 0.65 ± 0.10 | 0.69 ± 0.07 | 0.64 ± 0.09 | 0.63 ± 0.10 | 0.69 ± 0.07 | 0.90 ± 0.05 | 0.70 ± 0.09 | 0.10 ± 0.08 |
| Neoplastic Nuclear Features | 0.66 ± 0.07 | 0.69 ± 0.05 | 0.64 ± 0.06 | 0.63 ± 0.06 | 0.70 ± 0.05 | 0.87 ± 0.08 | 0.68 ± 0.04 | 0.08 ± 0.04 |
| Lesion & Epithelial Features | 0.67 ± 0.08 | 0.69 ± 0.05 | 0.65 ± 0.08 | 0.63 ± 0.07 | 0.71 ± 0.07 | 0.85 ± 0.09 | 0.69 ± 0.07 | 0.09 ± 0.07 |
| Clinical Features | 0.75 ± 0.05 | 0.75 ± 0.04 | 0.73 ± 0.05 | 0.78 ± 0.07 | 0.77 ± 0.05 | 0.86 ± 0.07 | 0.82 ± 0.07 | 0.22 ± 0.06 |
| Best Features per Theme | 0.75 ± 0.09 | 0.75 ± 0.08 | 0.74 ± 0.08 | 0.79 ± 0.08 | 0.80 ± 0.07 | 0.80 ± 0.14 | 0.85 ± 0.07 | 0.24 ± 0.07 |
| Top 10 Histological Features | 0.70 ± 0.09 | 0.71 ± 0.09 | 0.68 ± 0.08 | 0.70 ± 0.06 | 0.73 ± 0.07 | 0.81 ± 0.15 | 0.76 ± 0.06 | 0.16 ± 0.05 |
| Top 10 Features | **0.78 ± 0.05** | **0.79 ± 0.06** | **0.77 ± 0.07** | **0.82 ± 0.04** | **0.81 ± 0.08** | **0.86 ± 0.06** | **0.86 ± 0.06** | **0.26 ± 0.06** |

**Supplementary Table S2: Mean cross-validation performance metrics of the AutoGluon classifiers for the prediction of CIN2 lesion regression within two years.** The table summarizes the Weighted F1-score, Accuracy, Balanced Accuracy, AUROC, Precision, Recall, AUPRC, and AUPRC Above Baseline for models trained on various feature sets. Values are presented as mean ± standard deviation across all folds. The best metric value in each column is shown in bold. The Clinical Features demonstrate the highest performance among the independently evaluated feature groups (AUROC 0.78 ± 0.07), significantly outperforming all histological themes. However, the Top 10 Features (combined Clinical and Histological) consistently achieve the best overall performance across all metrics (AUROC 0.82 ± 0.04).

| Feature Set | F1 Weighted | Accuracy | Balanced Accuracy | AUROC | Precision | Recall | AUPRC | AUPRC Above Baseline |
|---|---|---|---|---|---|---|---|---|
| Immune Response Features | 0.64 ± 0.19 | 0.66 ± 0.17 | 0.61 ± 0.07 | 0.63 ± 0.07 | 0.52 ± 0.22 | 0.50 ± 0.19 | 0.44 ± 0.07 | 0.18 ± 0.07 |
| Neoplastic Nuclear Features | 0.40 ± 0.23 | 0.44 ± 0.19 | 0.54 ± 0.07 | 0.55 ± 0.07 | 0.32 ± 0.12 | **0.73 ± 0.20** | 0.38 ± 0.08 | 0.12 ± 0.07 |
| Lesion & Epithelial Features | 0.60 ± 0.14 | 0.60 ± 0.14 | 0.61 ± 0.05 | 0.64 ± 0.10 | 0.40 ± 0.11 | 0.61 ± 0.21 | 0.41 ± 0.10 | 0.15 ± 0.10 |
| Clinical Features | 0.77 ± 0.07 | 0.78 ± 0.08 | 0.69 ± 0.06 | 0.74 ± 0.09 | 0.66 ± 0.17 | 0.50 ± 0.10 | 0.59 ± 0.13 | 0.33 ± 0.13 |
| Best Features per Theme | 0.77 ± 0.05 | 0.77 ± 0.06 | 0.71 ± 0.04 | 0.76 ± 0.07 | 0.63 ± 0.14 | 0.59 ± 0.17 | 0.60 ± 0.09 | 0.34 ± 0.09 |
| Top 10 Histological Features | 0.64 ± 0.20 | 0.66 ± 0.19 | 0.61 ± 0.07 | 0.60 ± 0.10 | 0.55 ± 0.25 | 0.50 ± 0.23 | 0.47 ± 0.11 | 0.21 ± 0.11 |
| Top 10 Features | **0.81 ± 0.03** | **0.82 ± 0.04** | **0.73 ± 0.05** | **0.79 ± 0.07** | **0.74 ± 0.15** | 0.53 ± 0.14 | **0.65 ± 0.08** | **0.39 ± 0.09** |

**Supplementary Table S3: Mean cross-validation performance metrics of the AutoGluon classifiers model for the prediction of CIN2 regression within one year (quick regression)**. The table summarizes the Weighted F1-score, Accuracy, Balanced Accuracy, AUROC, Precision, Recall, AUPRC, and AUPRC Above Baseline for models trained on various feature sets. Values are presented as mean ± standard deviation across all folds. The best metric value in each column is shown in bold. The Clinical Features demonstrate the highest performance among the independently evaluated feature groups (AUROC 0.74 ± 0.09), with all histological themes showing substantially lower individual predictive capability. The Top 10 Features (combined Clinical and Histological) consistently achieve the best overall performance (AUROC 0.79 ± 0.07).

| **Model** | **Back bone** | **F1 Weighted** | **Accuracy** | **Balanced Accuracy** | **AURO C** | **Precisi on** | **Recall** | **AUP RC** | **AUPRC Above Baseline** |
|---|---|---|---|---|---|---|---|---|---|
| AttentionMIL | Uni | 0.50 ± 0.13 | 0.55 ± 0.09 | 0.55 ± 0.07 | 0.63 ± 0.09 | 0.64 ± 0.21 | 0.56 ± 0.32 | 0.74 ± 0.07 | 0.13 ± 0.08 |
| | Virch ow | 0.53 ± 0.08 | 0.58 ± 0.07 | 0.53 ± 0.07 | 0.60 ± 0.09 | 0.62 ± 0.05 | 0.78 ± 0.20 | 0.71 ± 0.07 | 0.11 ± 0.07 |
| Clam sb big | Uni | 0.61 ± 0.05 | 0.62 ± 0.06 | 0.60 ± 0.05 | 0.66 ± 0.08 | 0.68 ± 0.05 | 0.71 ± 0.16 | 0.76 ± 0.07 | 0.16 ± 0.07 |
| | Virch ow | 0.54 ± 0.08 | 0.58 ± 0.07 | 0.56 ± 0.08 | 0.65 ± 0.08 | 0.70 ± 0.14 | 0.64 ± 0.26 | 0.74 ± 0.07 | 0.14 ± 0.08 |
| Clam_sb small | Uni | 0.58 ± 0.08 | 0.60 ± 0.08 | 0.58 ± 0.07 | 0.65 ± 0.08 | 0.67 ± 0.05 | 0.67 ± 0.19 | 0.76 ± 0.07 | 0.16 ± 0.07 |
| | Virch ow | 0.57 ± 0.09 | 0.59 ± 0.08 | 0.59 ± 0.07 | 0.65 ± 0.08 | 0.68 ± 0.06 | 0.60 ± 0.23 | 0.74 ± 0.07 | 0.14 ± 0.07 |
| TransMIL | Uni | 0.50 ± 0.18 | 0.55 ± 0.12 | 0.57 ± 0.08 | 0.63 ± 0.08 | 0.55 ± 0.28 | 0.48 ± 0.31 | 0.74 ± 0.08 | 0.13 ± 0.08 |
| | Virch ow | 0.49 ± 0.15 | 0.54 ± 0.11 | 0.54 ± 0.09 | 0.62 ± 0.10 | 0.67 ± 0.24 | 0.55 ± 0.33 | 0.72 ± 0.07 | 0.12 ± 0.07 |
| Autogluon - Top 10 Features | **-** | **0.78 ± 0.05** | **0.79 ± 0.06** | **0.77 ± 0.07** | **0.82 ± 0.04** | **0.81 ± 0.08** | **0.86 ± 0.06** | **0.86 ± 0.06** | **0.26 ± 0.06** |

**Supplementary Table S4: Mean cross-validation performance metrics of MIL classifiers employing foundation-model backbone feature extractors for predicting two-year regression, presented alongside the results of our approach.** The table summarizes the Weighted F1-score, Accuracy, Balanced Accuracy, AUROC, Precision, Recall, AUPRC, and AUPRC Above Baseline for various MIL model configurations. Values are presented as mean ± standard deviation. The best metric value in each column is shown in bold. The AutoGluon model utilizing the Top 10 handcrafted features serves as a strong benchmark. The best performing MIL model, CLAM with the Uni backbone (AUROC 0.66 ± 0.08), is substantially outperformed by the AutoGluon Top 10 feature model (AUROC 0.82 ± 0.04) with over 16% increase in mean AUROC.

| Model | Back bone | F1 Weighted | Accuracy | Balanced Accuracy | AUR OC | Precision | Recall | AUPRC | AUPRC Above Baseline |
|---|---|---|---|---|---|---|---|---|---|
| AttentionMIL | Uni | 0.51 ± 0.13 | 0.54 ± 0.12 | 0.54 ± 0.07 | 0.60 ± 0.09 | 0.45 ± 0.26 | 0.55 ± 0.30 | 0.56 ± 0.20 | 0.12 ± 0.08 |
| | Virc how | 0.45 ± 0.22 | 0.48 ± 0.17 | 0.53 ± 0.08 | 0.59 ± 0.07 | 0.27 ± 0.10 | **0.63 ± 0.30** | 0.38 ± 0.09 | 0.12 ± 0.08 |
| Clam sb big | Uni | 0.60 ± 0.08 | 0.61 ± 0.09 | 0.57 ± 0.07 | 0.63 ± 0.09 | 0.51 ± 0.24 | 0.58 ± 0.29 | 0.59 ± 0.20 | 0.16 ± 0.09 |
| | Virc how | 0.59 ± 0.17 | 0.61 ± 0.15 | 0.56 ± 0.08 | 0.62 ± 0.07 | 0.33 ± 0.16 | 0.47 ± 0.28 | 0.45 ± 0.13 | 0.19 ± 0.12 |
| Clam sb small | Uni | 0.59 ± 0.10 | 0.61 ± 0.11 | 0.55 ± 0.08 | 0.60 ± 0.09 | 0.49 ± 0.22 | 0.50 ± 0.26 | 0.57 ± 0.20 | 0.14 ± 0.07 |
| | Virc how | 0.59 ± 0.15 | 0.61 ± 0.13 | 0.56 ± 0.05 | 0.64 ± 0.06 | 0.28 ± 0.15 | 0.45 ± 0.29 | 0.46 ± 0.09 | 0.19 ± 0.08 |
| TransMIL | Uni | 0.53 ± 0.19 | 0.58 ± 0.16 | 0.55 ± 0.07 | 0.60 ± 0.08 | 0.44 ± 0.31 | 0.41 ± 0.34 | 0.56 ± 0.19 | 0.13 ± 0.07 |
| | Virc how | 0.53 ± 0.18 | 0.58 ± 0.16 | 0.53 ± 0.08 | 0.59 ± 0.09 | 0.42 ± 0.29 | 0.37 ± 0.31 | 0.51 ± 0.19 | 0.13 ± 0.09 |
| Autogluon - Top 10 Features | - | **0.81 ± 0.03** | **0.82 ± 0.04** | **0.73 ± 0.05** | **0.79 ± 0.07** | **0.74 ± 0.15** | 0.53 ± 0.14 | **0.65 ± 0.08** | **0.39 ± 0.09** |

**Supplementary Table S5: Mean cross-validation performance metrics of MIL classifiers employing foundation-model backbone feature extractors for predicting one-year regression (quick regression), presented alongside the results of our approach.** The table summarizes the Weighted F1-score, Accuracy, Balanced Accuracy, AUROC, Precision, Recall, AUPRC, and AUPRC Above Baseline for various MIL model configurations. Values are presented as mean ± standard deviation. The best metric value in each column is shown in bold. The AutoGluon model utilizing the Top 10 handcrafted features performs significantly better compared to MIL models. The best performing MIL model, CLAM with the Uni backbone (AUROC 0.64 ± 0.06), is substantially outperformed by the AutoGluon Top 10 feature model (AUROC 0.79 ± 0.07) with over 15% increase in mean AUROC.
